# RNAseq-Based Machine Learning Models for Prognostication of Multiple Myeloma

**DOI:** 10.1101/2025.01.31.25321495

**Authors:** Krish U. Shah, Kaina A. Millan, Anna E. Pula, Tadeusz F. Kubicki, Joseph Cannova, Sulin Wu, Manoj Bhagwat, Quincy C. Guenther, Jennifer Cooperrider, Gregory Roloff, Aarti Venkat, Benjamin A. Derman, Andrzej J. Jakubowiak, Michael W. Drazer

**Affiliations:** Section of Hematology/Oncology, University of Chicago, IL, USA; Department of Hematology, Medical University of Lodz, Poland; Section of Biomedical Data Science, University of Chicago, Chicago, IL, US; Section of Genetic Medicine, University of Chicago, Chicago, IL, USA

**Author notes:** **Author Approval:** This manuscript has been seen and approved by all listed authors. This manuscript has not been accepted or published by a journal. **Competing Interests: TFK:** honoraria (Jannsen) **AEP:** honoraria (Jannsen, Roche, Amgen); clinical trial support (Sanofi, GSK) **BAD:** consulting or advisory roles (Johnson and Johnson, Sanofi, Canopy, COTA); research funding (Amgen, GSK); honoraria (Multiple Myeloma Research Foundation); independent reviewer for clinical trial (BMS) **AJJ:** consulting or advisory roles (Abbvie, Amgen, BMS, Celgene, GSK, Gracell, Janssen, Karyopharm Therapeutics, Sanofi) **MWD:** consulting or advisory roles (Argenx); honoraria for educational writing (American Society of Hematology Self-Assessment Program); honoraria (Novartis). **Data Declarations:** Clinical, biochemical, survival, and RNAseq data was acquired from the semi-public Multiple Myeloma Research Foundation (MMRF) Research Gateway (https://research.themmrf.org/). Raw whole exome sequencing data was downloaded from the Database of Genotype and Phenotype (#phs000748.v7.p4, project #34982, https://www.ncbi.nlm.nih.gov/projects/gap/cgi-bin/study.cgi?study_id=phs000748.v7.p4). **Data Availability:** All data produced in the present study are available upon reasonable request to the authors. **Code Availability:** The code for pre-processing the data, training, and validating the models can be requested by email to the corresponding author. **Funding Statement:** This study did not receive any funding.

## Abstract

**Background:** Multiple myeloma (MM) is characterized by abnormal plasma cell proliferation in the bone marrow, leading to symptoms like osteolytic lesions, anemia, hypercalcemia, and elevated serum creatinine. RNA-sequencing-based prognostic indicators for MM have shown promise in stratifying risk and assessing first-line treatment options. This study uses machine learning techniques and leverages RNA-sequencing, clinical, and biochemical data from the Multiple Myeloma Research Foundation (MMRF) CoMMpass cohort to predict patient prognosis.

**Methods:** RNAseq data of 60,623 genes from bone marrow samples of 708 MM patients were pre-processed for batch effect correction and split into training (70%) and testing (30%) sets. Feature selection involved MAD, mRMR, and iterative permutation importance filtering for predicting PFS and OS. Machine learning survival models like Random Survival Forest (RSF), Gradient Boosted (GB), and Component-wise Gradient Boosted (CGB) were developed and optimized. Performance was evaluated using C-index and integrated Brier score (IBS).

**Results:** The RSF and GB models showed the highest performance for predicting progression-free survival (PFS) and overall survival (OS) on the testing dataset. Significant features for PFS included stem cell transplant status, serum β2-microglobulin levels, germline mutational status, and expression of *C12orf75* and ENSG00000256006. For OS, stem cell transplant status, age, serum β2-microglobulin levels, germline mutational status, and expression of *NUTM2B-AS1* and ENSG00000287022 were prominent. Gene ontology analyses confirmed the biological relevance of enriched pathways related to cell division, protein localization, and cancer.

**Conclusion:** Integrating RNAseq and clinical data with advanced machine learning models presents a robust approach for predicting MM prognosis, highlighting gene expression programs, germline mutational status, and clinical markers as significant features. Future research should focus on independent validation to confirm findings and explore additional genomic data for enhanced prognostication.

---

Multiple myeloma (MM) is characterized by the clonal proliferation of abnormal plasma cells in the bone marrow (Cowan et al., 2022). Osteolytic lesions and anemia are the most common findings among patients with MM, closely followed by hypercalcemia and elevated serum creatinine (Rajkumar & Kumar, 2016). These markers have been adopted into diagnostic criteria for MM, which, alongside molecular cytogenetic classification, have been consolidated into the International Staging System (ISS) for risk stratification (Rajkumar & Kumar, 2016). Chromosome ploidy and translocations involving oncogenes significantly associate with downstream patient prognosis and underscore the importance of cytogenetic classification for treatment and follow-up protocols (Sawyer, 2011). Broadly, there are two karyotypic groups: hyperdiploid (HRD) and non-HRD phenotypes, the latter of which generally exhibit other genetic aberrations such as chromosome trisomies, immunoglobulin translocations, and copy number alterations. These abnormalities ultimately alter gene expression and allow for the characterization of RNA-sequencing molecular subtypes of MM (Skerget et al., 2024).

RNA expression patterns in MM are significantly associated with copy number and immunoglobulin translocations, thereby conferring a prognostic signature without whole-genome or whole-exome sequencing (Skerget et al., 2024). Before Skerget and colleagues’ analysis of the Multiple Myeloma Research Foundation (MMRF) CoMMpass dataset, groups had proposed the utility of RNA-sequencing-based prognostic indicators in better stratifying risk and assessing first-line treatment options (Zamani-Ahmadmahmudi et al., 2020; Emde-Rajaratnam et al., 2023). Our study leverages the MMRF CoMMpass cohort’s RNA-sequencing, clinical, and biochemical data to predict patient prognosis using machine learning techniques. To our knowledge, this study is also the first to incorporate germline mutational data into a MM-focused machine learning model.

While a prior MM machine learning study used a random forest model to predict overall survival (OS) (Orgueira et al., 2021), our study adds to these prior findings by also developing Gradient Boosted (GB) and Component-wise Gradient Boosted (CGB) models that incorporate germline mutational data and predict both patient progression-free survival (PFS) and OS. Studying PFS and OS in MM is particularly relevant, as PFS is less likely to be biased by attrition and confounding effects from crossover/post-progression therapies when compared with OS (Cartier et al., 2015; Holstein et al., 2019). Furthermore, the CoMMpass dataset was primarily generated from newly diagnosed and untreated MM patients, which makes it particularly well-suited for studies of PFS. By evaluating the performance of various machine learning models and feature selection techniques, we identified features for predicting PFS and OS in MM patients.

## METHODS

### Participants

We utilized de-identified and processed RNAseq count data for participants involved in the MMRF CoMMpass Interim Analysis 22. Sample collection, storage, RNAseq library preparation, and sequencing have been described previously (Skerget et al., 2024). RNAseq quality control metrics and clinical, biochemical, and survival data were obtained from the MMRF Research Gateway and supplementary information provided by Skerget and colleagues (Skerget et al., 2024).

### Germline Mutational Status

The GVCF workflow by GATK4.4 and HaplotypeCaller were applied to raw whole exome sequencing data downloaded from the Database of Genotype and Phenotype (#phs000748.v7.p4, project #34982). Called germline variants were filtered and functionally annotated using Funcotator. Variants associated with hereditary cancer, hematopoietic malignancies, bone marrow failure, telomere biology, and/or immunodeficiency syndromes were analyzed alongside candidate genes for hereditary MM (Catalano et al., 2021). Pathogenicity was determined by consulting the ClinVar database and/or guidelines from the American College of Medical Genetics and Genomics.

### Pre-Processing

Participants for whom RNAseq data passed publication quality control were included (Skerget et al., 2024). Before feature selection and downstream analyses, the expression data of 60,623 genes and 708 bone marrow tumor samples were corrected for confounding biobank batch effects using the R packages DESeq2 and Limma. Principal component analysis (PCA) was used as a validation tool before and after batch effect correction. Following batch effect correction, the data was split 70/30 into training (n=496) and testing (n=212) sets.

### Feature Selection

Several techniques for feature selection were attempted, including but not limited to variance thresholding, maximum relevance minimum redundancy (mRMR) selection, Lasso Cox regression, single gene clusterization, and dimensionality reduction. Each feature selection strategy involved imputing missing clinical and biochemical variables using the impute function from the randomForestSRC R package and the associated default parameters. Little’s Missing Completely at Random (MCAR) test was used to justify imputation. After assessing the concordance index (C-index) on the holdout testing dataset, the most optimal feature selection strategy was selected. This strategy involved choosing the top 30,000 most variable log-transformed and centered genes using median absolute deviation (MAD) and then applying mRMR on the geneset using the mRMRe R package. The top 100 most relevant genes correlated with PFS and OS were combined with the imputed clinical and biochemical features. One hundred genes were chosen as an endpoint for the mRMR algorithm as testing various feature selection techniques suggested an optimal dimensionality of between 30 and 80 features. These 100 features were then used for survival analysis, and each machine learning model was further refined by excluding features whose permutation importance, after adding one standard deviation, was less than or equal to zero. The machine learning workflow is visually represented in **Figure 1**.

**Figure 1.**
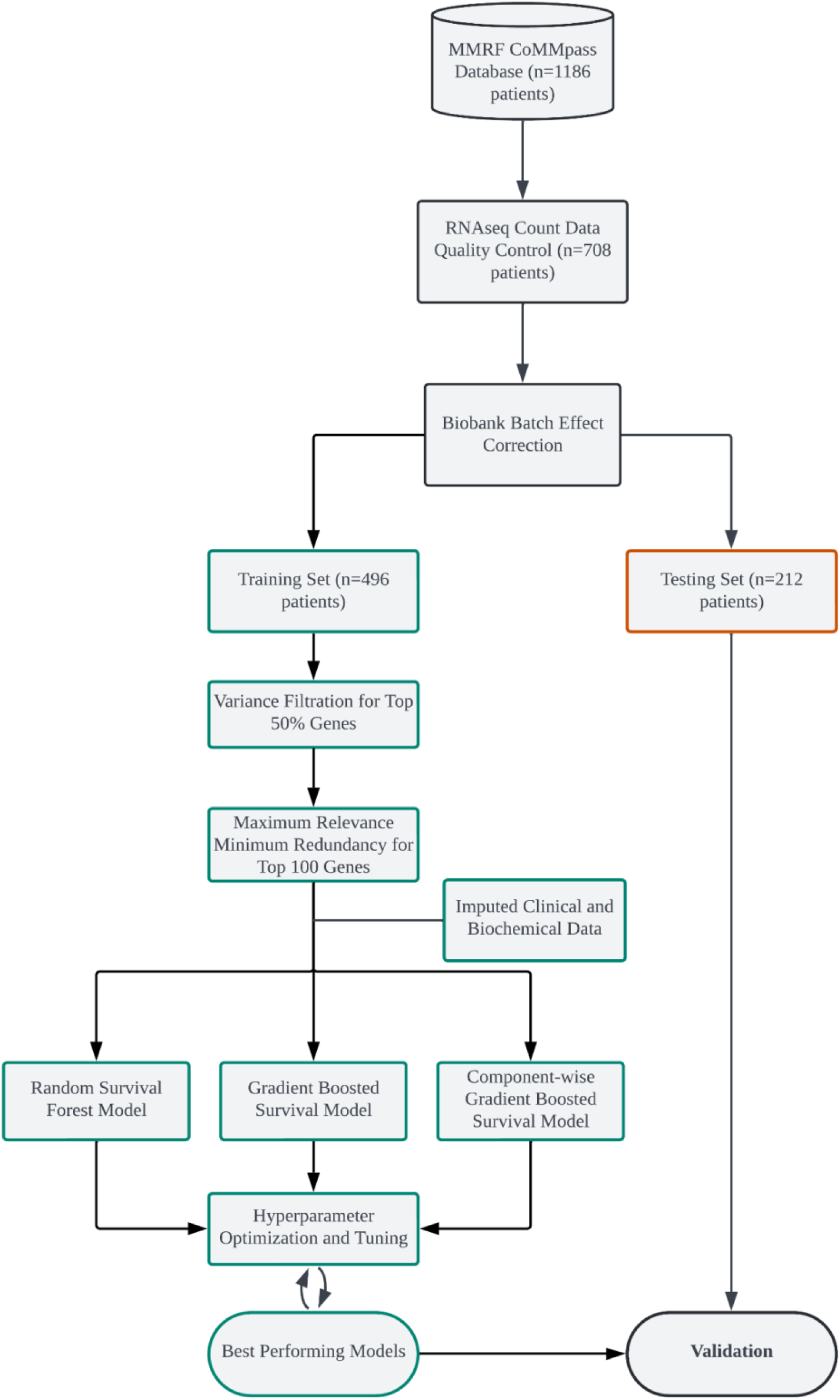
Machine Learning Workflow *Note.* Variance calculated using median absolute deviation (MAD). Each model was independently optimized using Optuna and tuned by iteratively removing variables with mean importance plus one standard deviation less than or equal to zero.

### Geneset Enrichment and Ontology

The top 100 most relevant genes for each outcome (PFS and OS) served as input to the Metascape web portal (Zhou et al., 2019). Significantly enriched biological pathways were visualized using the “Express Analysis” option.

### Survival Analysis

Machine learning survival models were developed and tuned in Python. Random Survival Forest (RSF), Gradient Boosted (GB), and Component-wise Gradient Boosted (CGB) models were developed using the scikit-survival Python package. Hyperparameters for the models were tuned using the Optuna Python package maximizing C-index and the integrated Brier score (IBS) over five folds of Monte-Carlo cross-validation using an 80/20 training to validation split. Fifty optimization trials using a Tree-structured Parzen Estimator (TPE) sampler and a median pruner were utilized for hyperparameter tuning. Hyperparameter grids were tuned by visualizing performance and importance using Optuna visualizations. Each optimized model was assessed on the testing dataset, and the C-index and IBS metrics were obtained.

### Variable Importances

The permutation importance function with 30 repeats from the scikit-learn Python package was used to derive variable importances for all models.

## RESULTS

### Gene Ontology Enrichment

The PFS geneset (n=100) was significantly enriched for the following biological pathways and associated terms: negative regulation of protein localizations (GO:1903828), determination of left/right symmetry (GO:0007368), cell morphogenesis (GO:0000902), integrated breast cancer pathway (WP1984), cell division (GO:0051301), epidermis development (GO:0008544), vacuolar transport (GO:0007034), and negative regulation of gene expression, epigenetic (GO:0045814) **(Figure 2)**.

**Figure 2.**
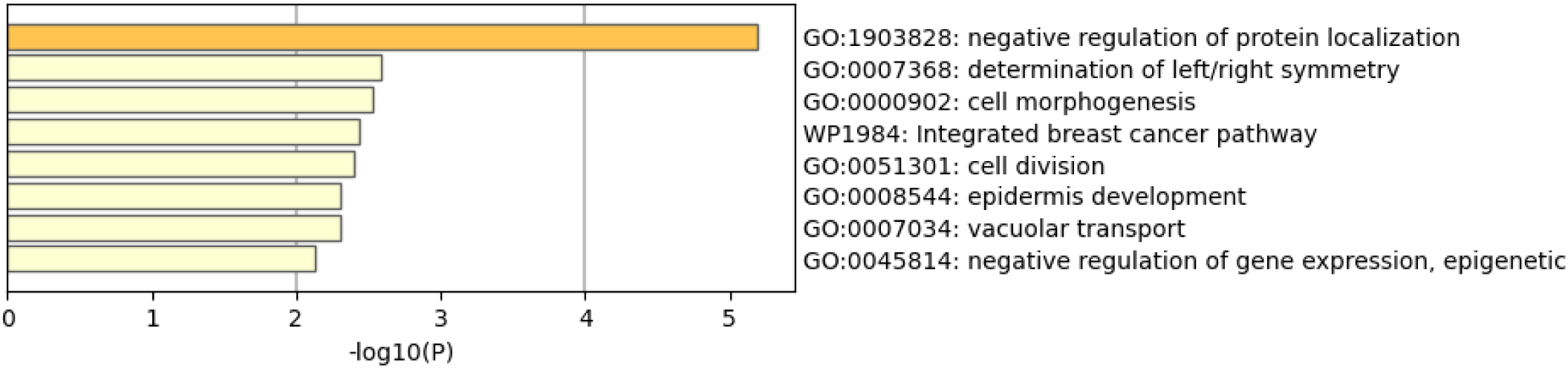
GO Term Enrichment Analysis for PFS Geneset (n=100)

The OS geneset (n=100) was significantly enriched for the following biological pathways and associated terms: Wnt signaling pathway (GO:0016055), RNA splicing via transesterification reactions with bulged adenosine as nucleophile (GO:0000377), PPARA activates gene expression (R-HSA-1989781), cell population proliferation (GO:0008283), negative regulation of neuron projection development (GO:0010977), mitotic prophase (R-HSA- 68875), and cellular response to organic cyclic compound (GO:0071407) **(Figure 3)**.

**Figure 3.**
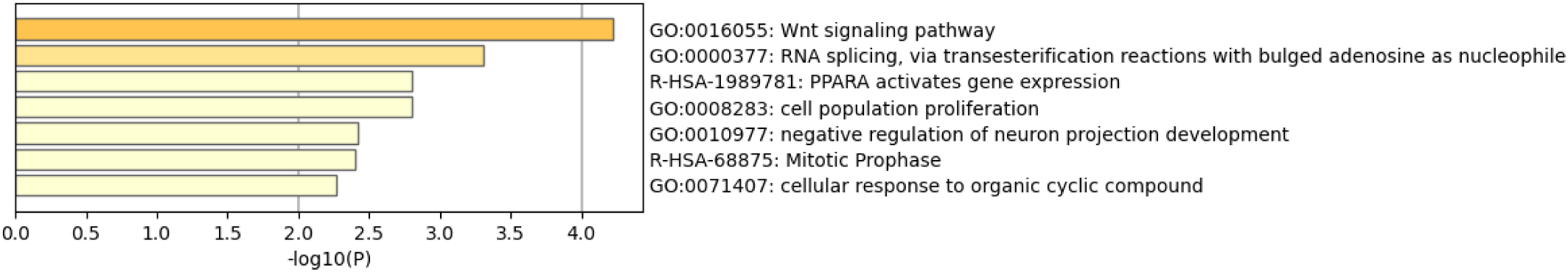
GO Term Enrichment Analysis for OS Geneset (n=100)

**Figure 3.**
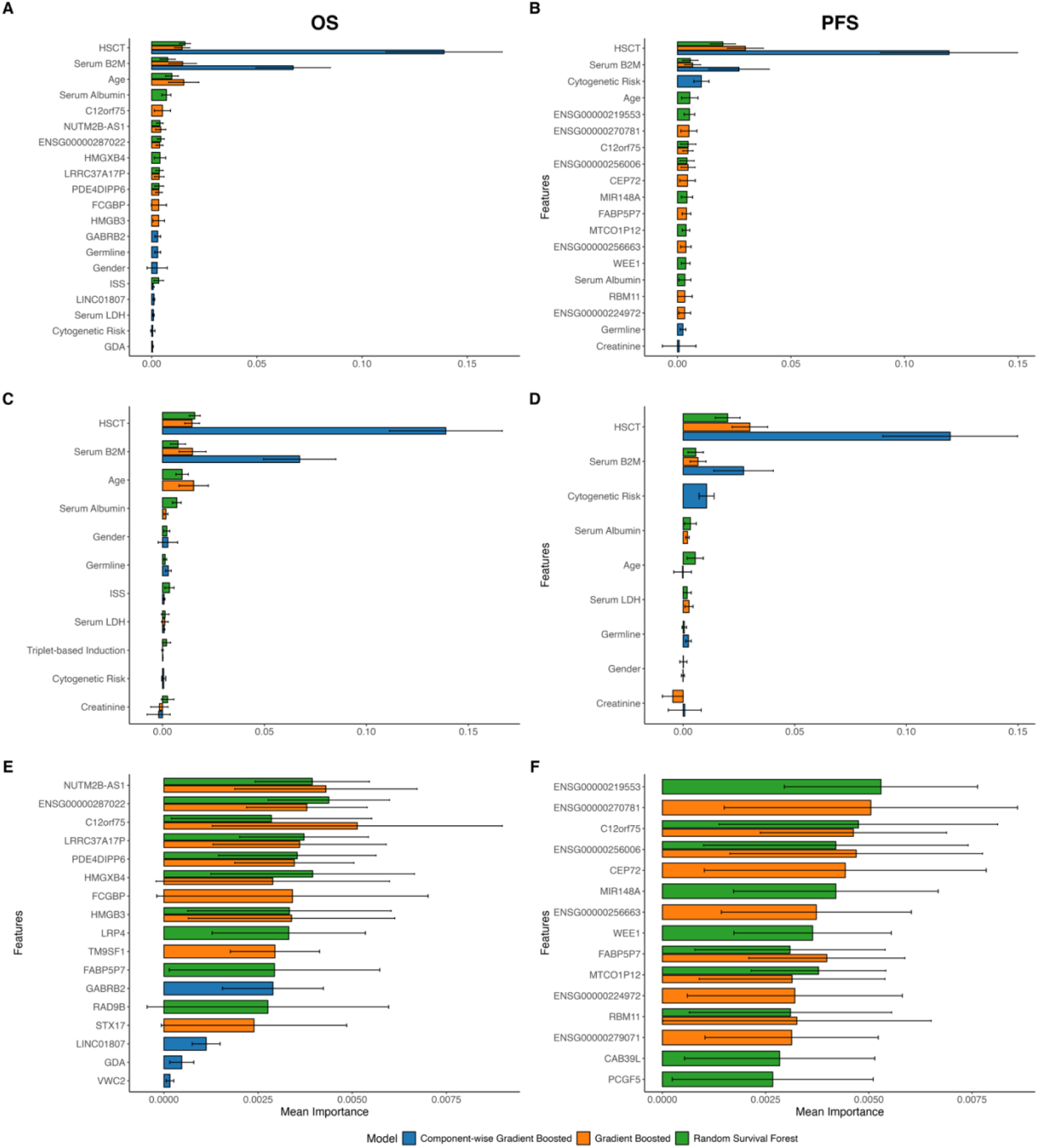
Top Total, Gene Expression, and Clinical Features by Model for Each Outcome **A** Top 10 total features by model for PFS. Error bars represent mean importance plus or minus the associated standard deviation. **B** Top 10 total features by model for OS. **C** Top 10 clinical features by model for PFS. **D** Top 10 total features by model for OS. **E** Top 10 gene expression features by model for PFS. **F** Top 10 gene expression features by model for OS.

### PFS

Neither PFS nor the clinical variables differed significantly between the training and testing datasets **(Table 1)**. Performance on the training dataset was the highest for the GB model (RSF: 0.749; GB: 0.818; CGB: 0.695) **(Table 2)**. Performance on the testing dataset was the highest for the RSF model for C-index (RSF: 0.746; GB: 0.730; CGB: 0.678) and the GB model for IBS (RSF: 0.164; GB: 0.158; CGB: 0.176) **(Table 2)**. Hematopoietic stem cell transplant (HSCT) status, serum β2-microglobulin levels, age, ENSG00000219553 gene expression, and *C12orf75* gene expression were the most significant RSF features for predicting PFS **(Table 3)**. HSCT status, serum β2-microglobulin levels, ENSG00000270781 gene expression, ENSG00000256006 gene expression, and *C12orf75* gene expression were the most significant GB features for predicting PFS **(Table 4**). HSCT status, serum β2-microglobulin levels, cytogenetic risk, germline mutational status, and creatinine were the most significant CGB features for predicting PFS **(Table 5)**.

**Table 1.** Descriptors of Clinical Variables.

|  |  | Training | Testing | *P* values |
| --- | --- | --- | --- | --- |
| **N** |  | 496 | 212 |  |
| **PFS (days), median [IQR]** |  | 715.0 [315.8, 1547.5] | 660.0 [237.0, 1533.0] | 0.29 |
| **PFS, n (%)** | 0 | 201 (40.5) | 93 (43.9) | 0.46 |
|  | 1 | 295 (59.5) | 119 (56.1) |  |
| **OS (days), median [IQR]** |  | 1575.0 [415.3, 2576.3] | 1390.5 [417.5, 2598.8] | 0.61 |
| **OS, n (%)** | 0 | 321 (64.7) | 145 (68.4) | 0.39 |
|  | 1 | 175 (35.3) | 67 (31.6) |  |
| **Age (years), mean (SD)** |  | 62.7 (10.5) | 62.0 (11.4) | 0.45 |
| **Cytogenetic Risk, n (%)** | 0 | 160 (32.3) | 67 (31.6) | 0.31 |
|  | 1 | 273 (55.0) | 109 (51.4) |  |
|  | NA | 63 (12.7) | 36 (17.0) |  |
| **Creatinine (μmol/L), median [IQR]** |  | 88.4 [70.7, 118.5] | 92.8 [68.7, 115.1] | 0.82 |
| **Triplet-based induction, n (%)** | 0 | 120 (24.2) | 47 (22.2) | 0.48 |
|  | 1 | 361 (72.8) | 155 (73.1) |  |
|  | NA | 15 (3.0) | 10 (4.7) |  |
| **HSCT, n (%)** | 0 | 240 (48.4) | 101 (47.6) | 0.54 |
|  | 1 | 241 (48.6) | 101 (47.6) |  |
|  | NA | 15 (3.0) | 10 (4.7) |  |
| **ISS, n (%)** | 1 | 170 (34.3) | 76 (35.8) | 0.26 |
|  | 2 | 184 (37.1) | 64 (30.2) |  |
|  | 3 | 132 (26.6) | 65 (30.7) |  |
|  | NA | 10 (2.0) | 7 (3.3) |  |
| **Gender, n (%)** | 0 | 292 (58.9) | 126 (59.4) | 0.96 |
|  | 1 | 204 (41.1) | 86 (40.6) |  |
| **Germline, n (%)** | 0 | 414 (83.5) | 178 (84.0) | 0.41 |
|  | 1 | 67 (13.5) | 24 (11.3) |  |
|  | NA | 15 (3.0) | 10 (4.7) |  |
| **Serum β2M (mcg/mL), median [IQR]** |  | 3.7 [2.5, 5.8] | 3.5 [2.6, 6.5] | 0.47 |
| **Serum Albumin (g/L), median [IQR]** |  | 36.0 [32.0, 40.0] | 37.0 [33.0, 41.9] | 0.18 |
| **Serum LDH (μkat/L), median [IQR]** |  | 2.9 [2.3, 4.0] | 2.8 [2.3, 3.6] | 0.24 |

*Note.* Cytogenetic risk is defined as: del(17p), amp(1q), del(1p), t(4;14), t(14;16), or t(14;20). PFS 0 is defined as no progression or right censored and PFS 1 is defined as progression. OS 0 is defined as alive or right censored and OS 1 is defined as death. Gender 0 is defined as Male and Gender 1 is defined as Female.

**Table 2.** Model Results.

|  | Training |  | Testing |  |  |  |
| --- | --- | --- | --- | --- | --- | --- |
|  | PFS | OS | PFS |  | OS |  |
|  | C-Index | C-Index | C-Index | Integrated Brier Score | C-Index | Integrated Brier Score |
| Random Survival Forest | 0.749 | 0.798 | 0.746 | 0.164 | 0.800 | 0.142 |
| Gradient Boosted | 0.818 | 0.886 | 0.730 | 0.158 | 0.801 | 0.141 |
| Component-wise Gradient Boosted | 0.695 | 0.710 | 0.678 | 0.176 | 0.736 | 0.160 |

**Table 3.**
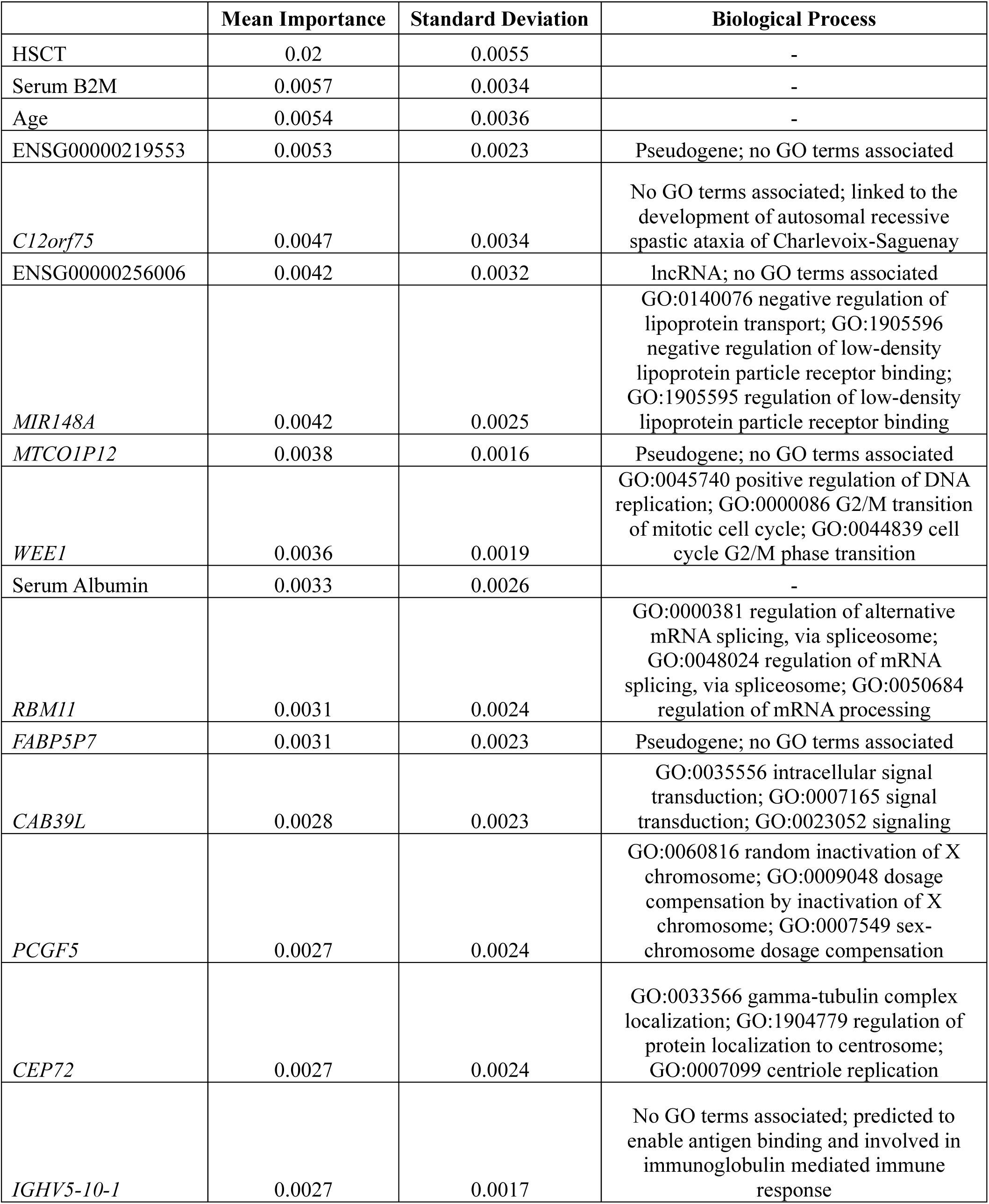
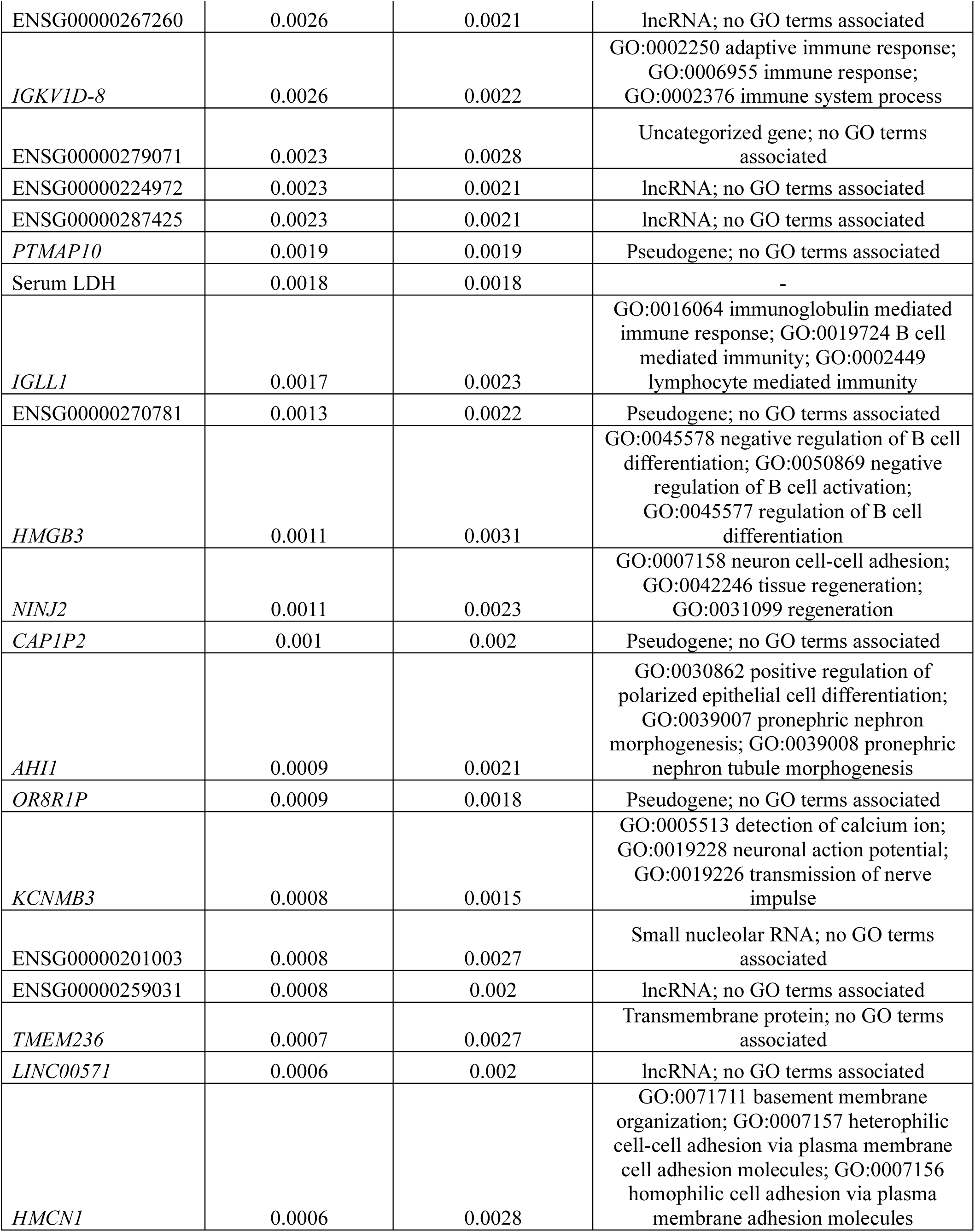
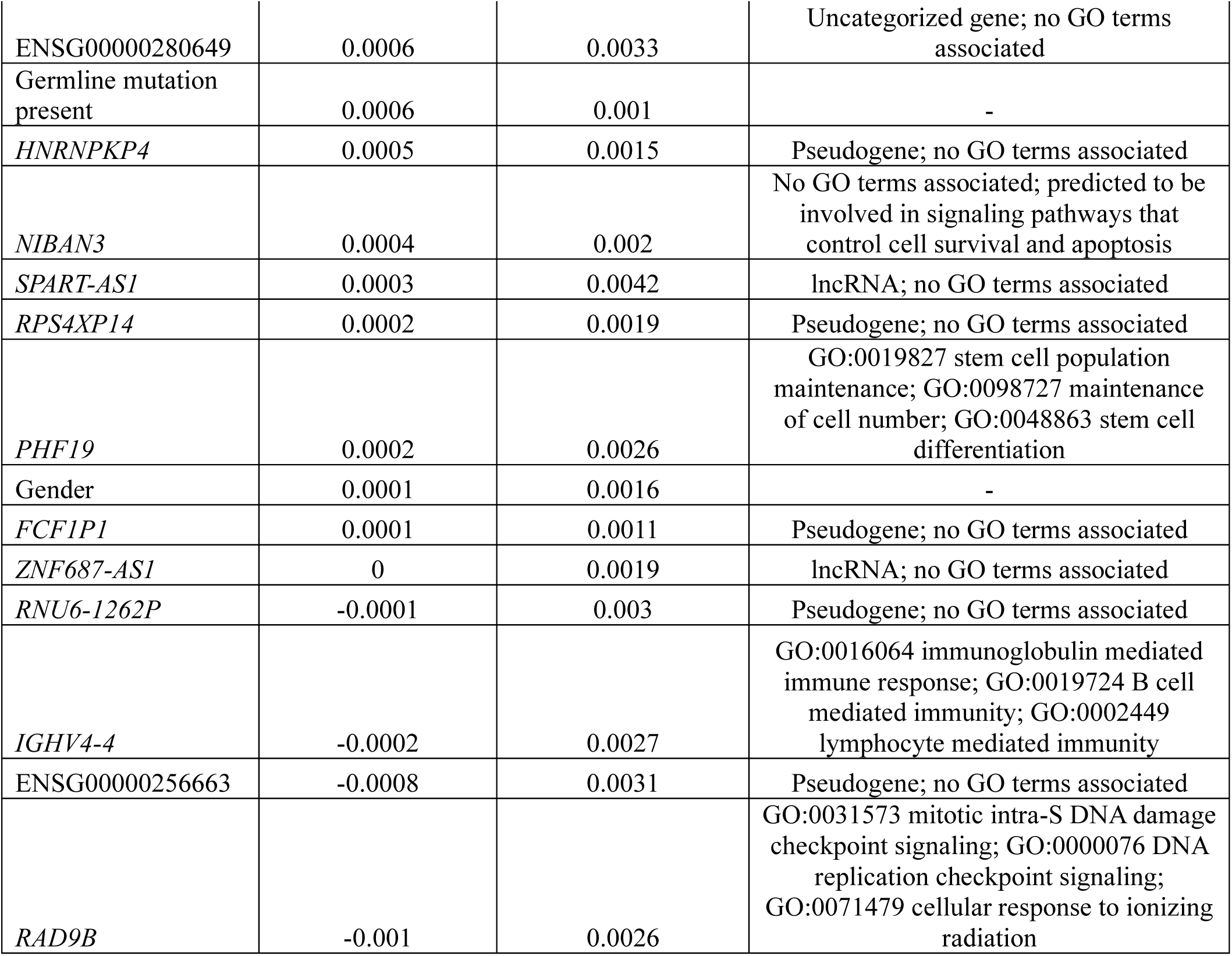
Random Survival Forest Variable Importances for PFS.

**Table 4.**
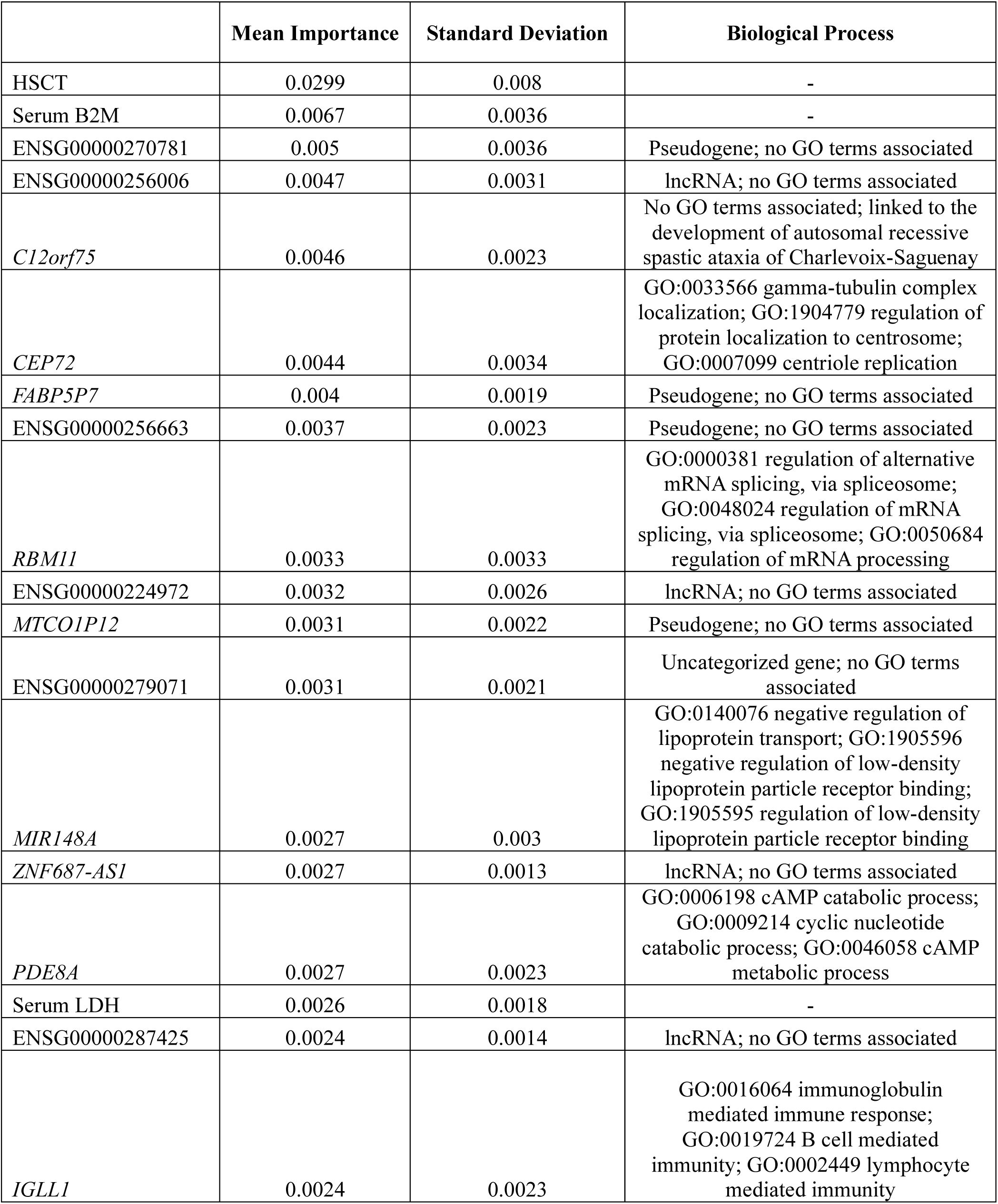
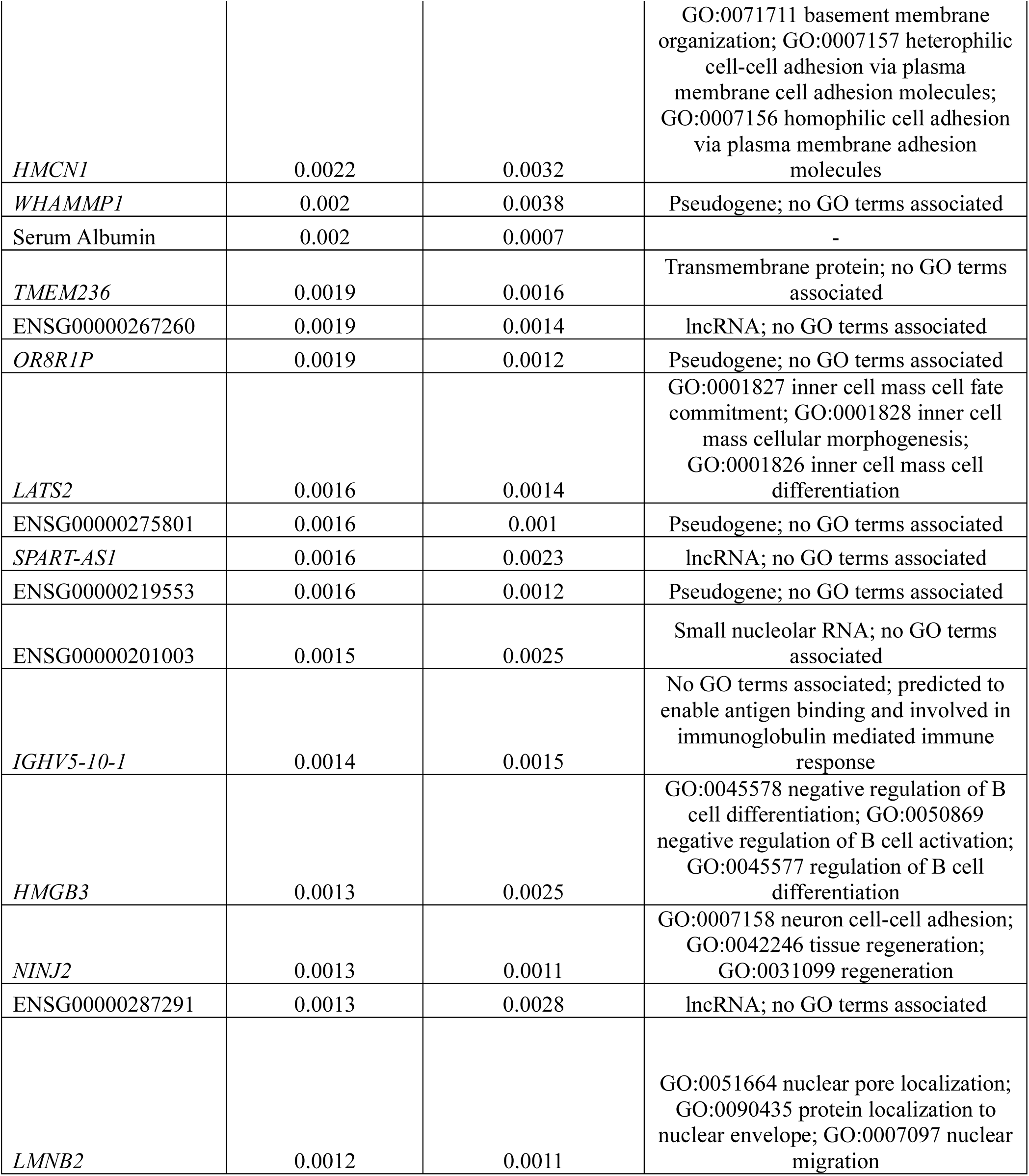
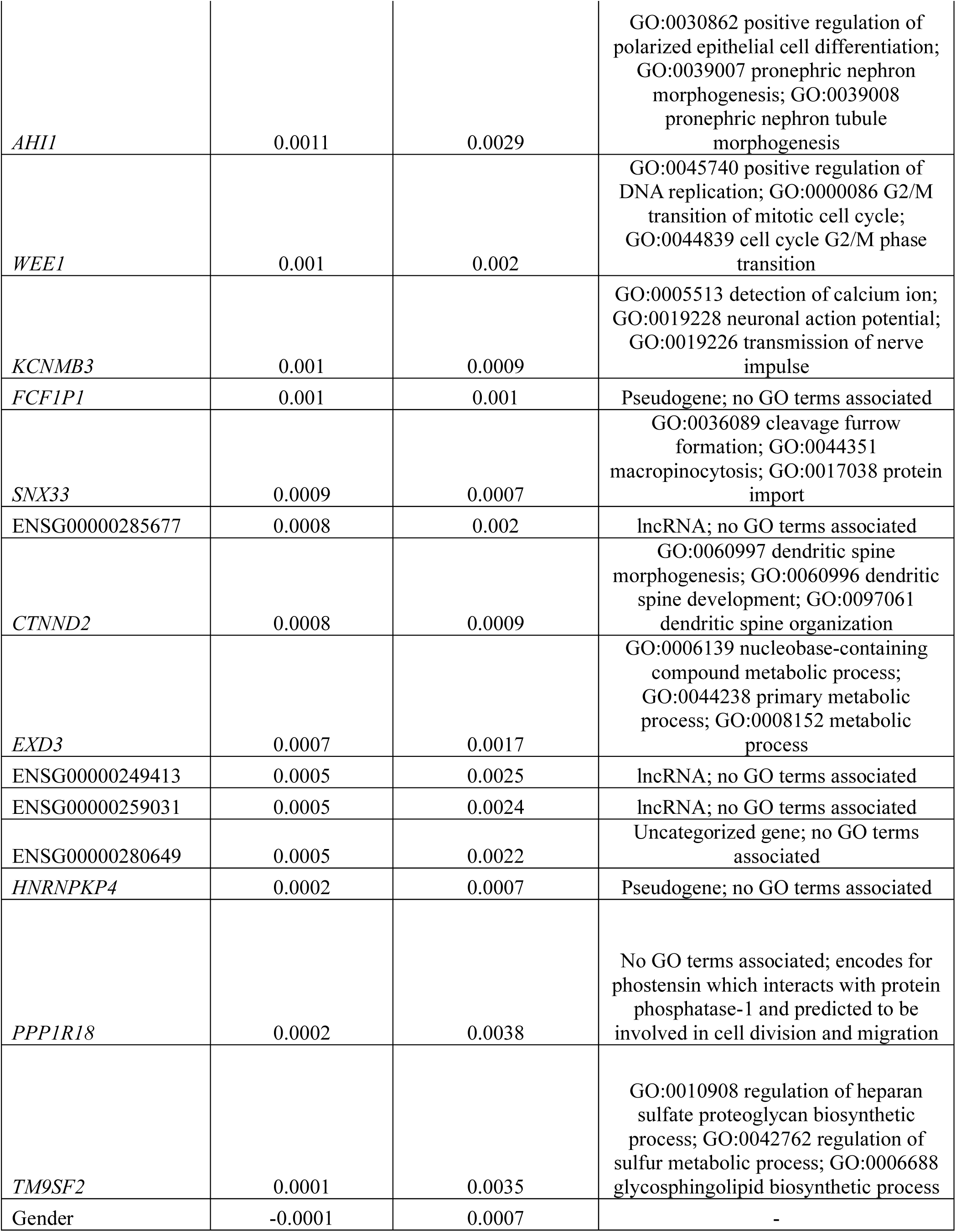
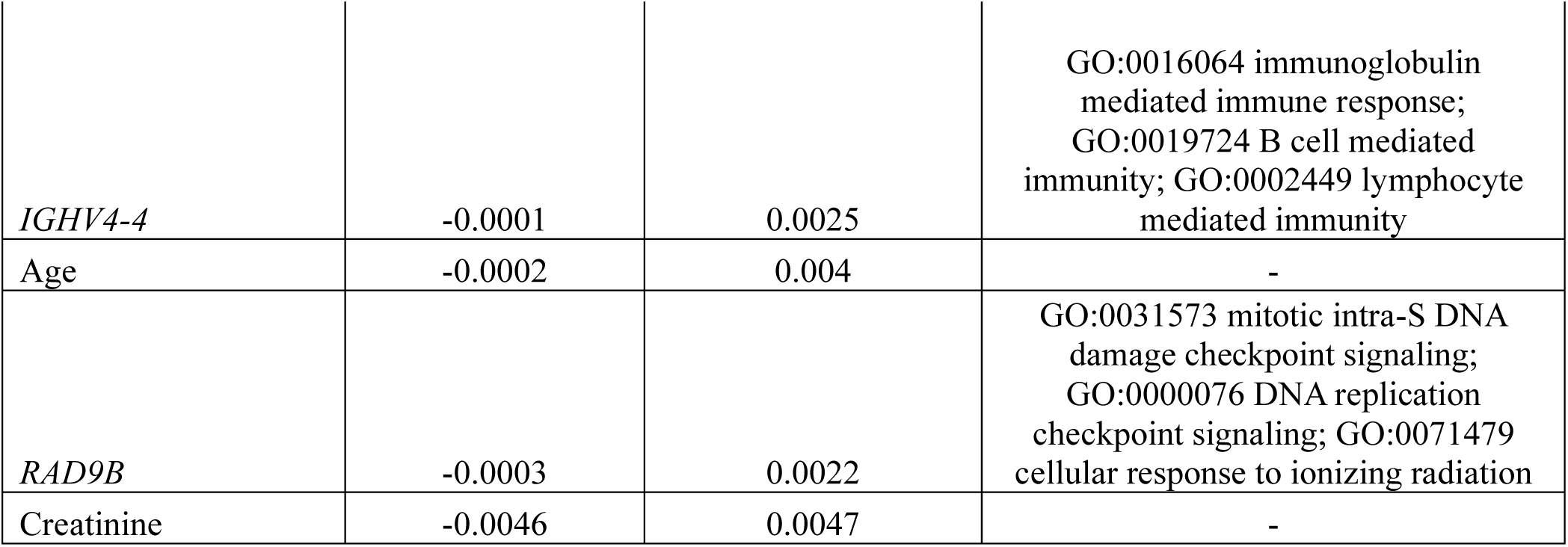
Gradient Boosted Variable Importances for PFS.

**Table 5.** Component-wise Gradient Boosted Variable Importances for PFS.

|  | **Mean Importance** | **Standard Deviation** | **Biological Process** |
| --- | --- | --- | --- |
| HSCT | 0.1196 | 0.0302 | - |
| Serum B2M | 0.0271 | 0.0133 | - |
| Cytogenetic Risk | 0.0105 | 0.0033 | - |
| Germline mutation present | 0.0024 | 0.0012 | - |
| Creatinine | 0.0008 | 0.0074 | - |

### OS

Neither OS nor the clinical variables differed significantly between the training and testing datasets. Out-of-bag performance on the training dataset was the highest for the GB model (RSF: 0.798; GB: 0.886; CGB: 0.710) **(Table 2)**. Similar patterns were observed for performance on the testing dataset for C-index (RSF: 0.800; GB: 0.801; CGB: 0.736) **(Table 2)** and IBS (RSF: 0.142; GB: 0.141; CGB: 0.160). HSCT status, age, serum β2-microglobulin levels, serum albumin levels, and ENSG00000287022 gene expression were the most significant RSF features for predicting OS **(Table 6)**. Age, serum β2-microglobulin levels, HSCT status, *C12orf75* gene expression, and *NUTM2B-AS1* gene expression were the most significant GB features for predicting OS **(Table 7)**. HSCT status, serum β2-microglobulin levels, *GABRB2* gene expression, germline mutational status, and gender were the most significant CGB features for predicting OS **(Table 8)**.

**Table 6.**
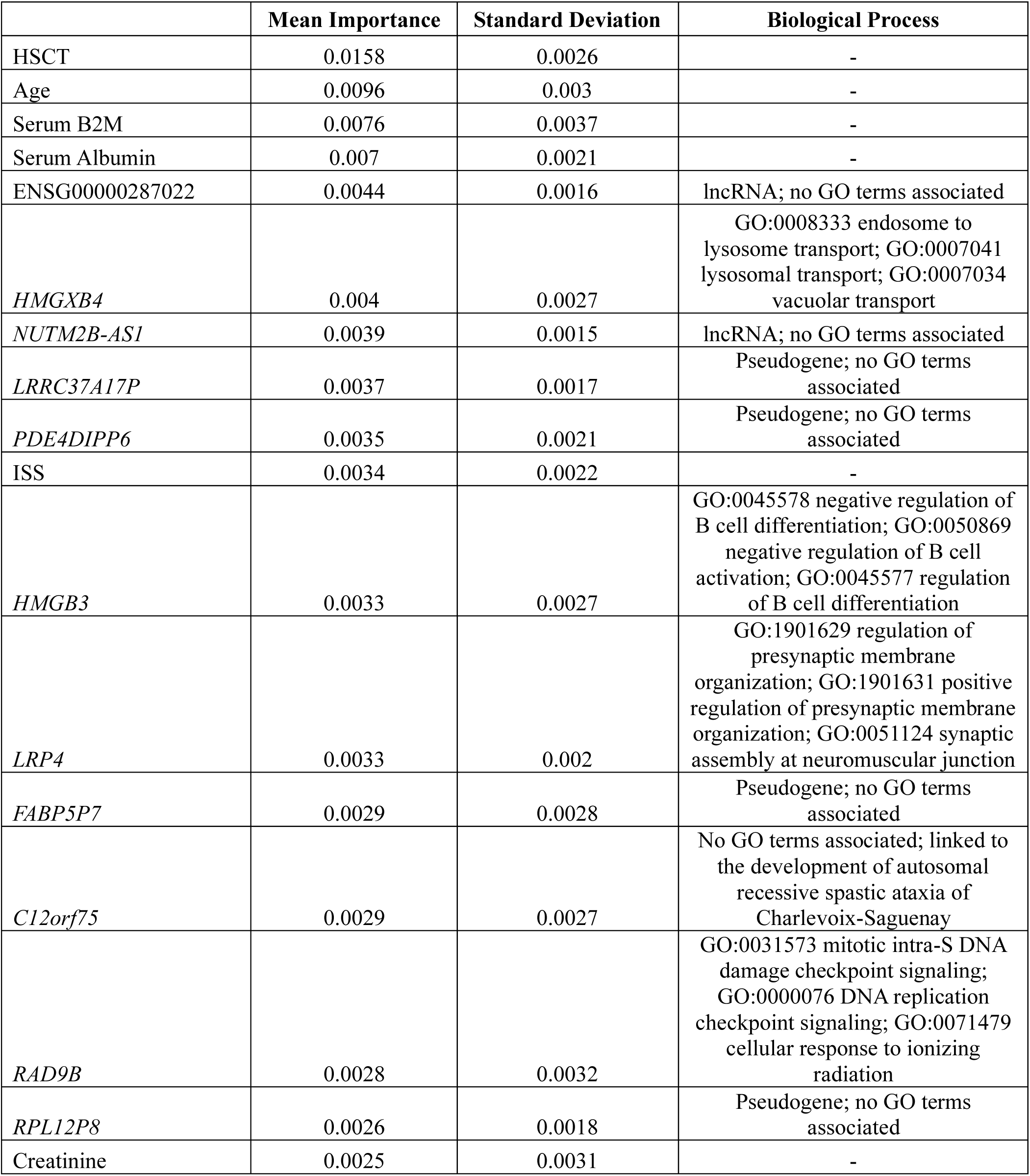
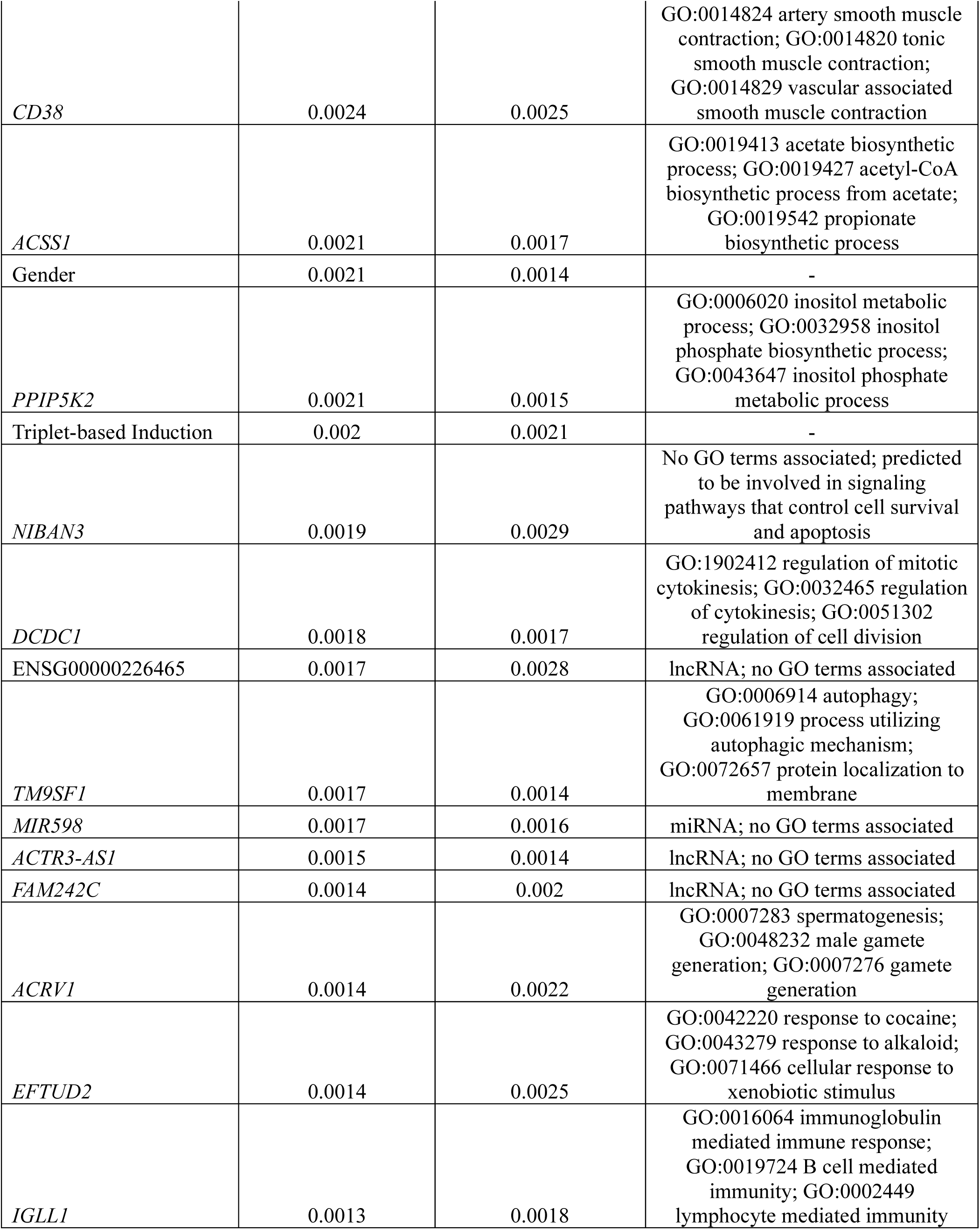
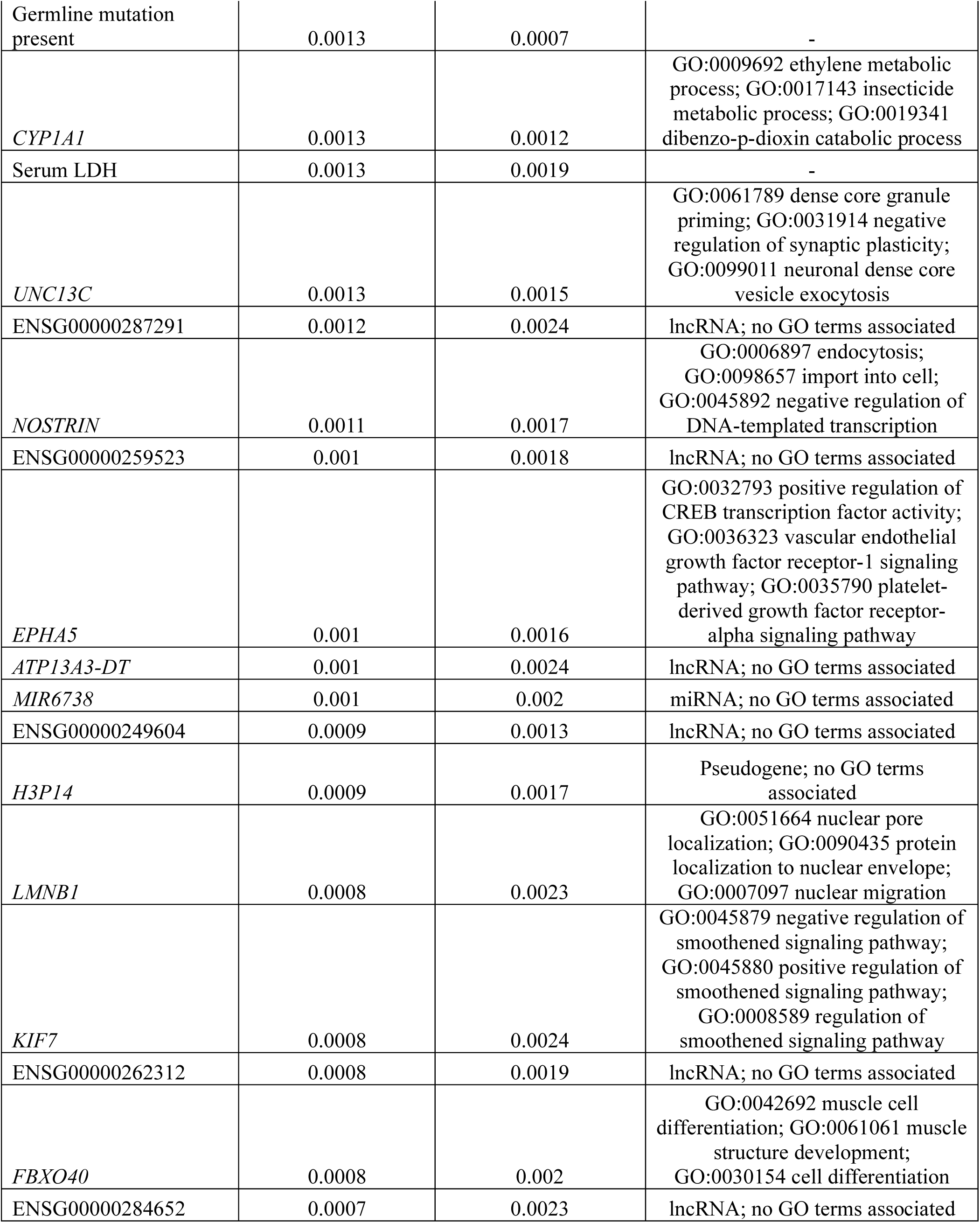
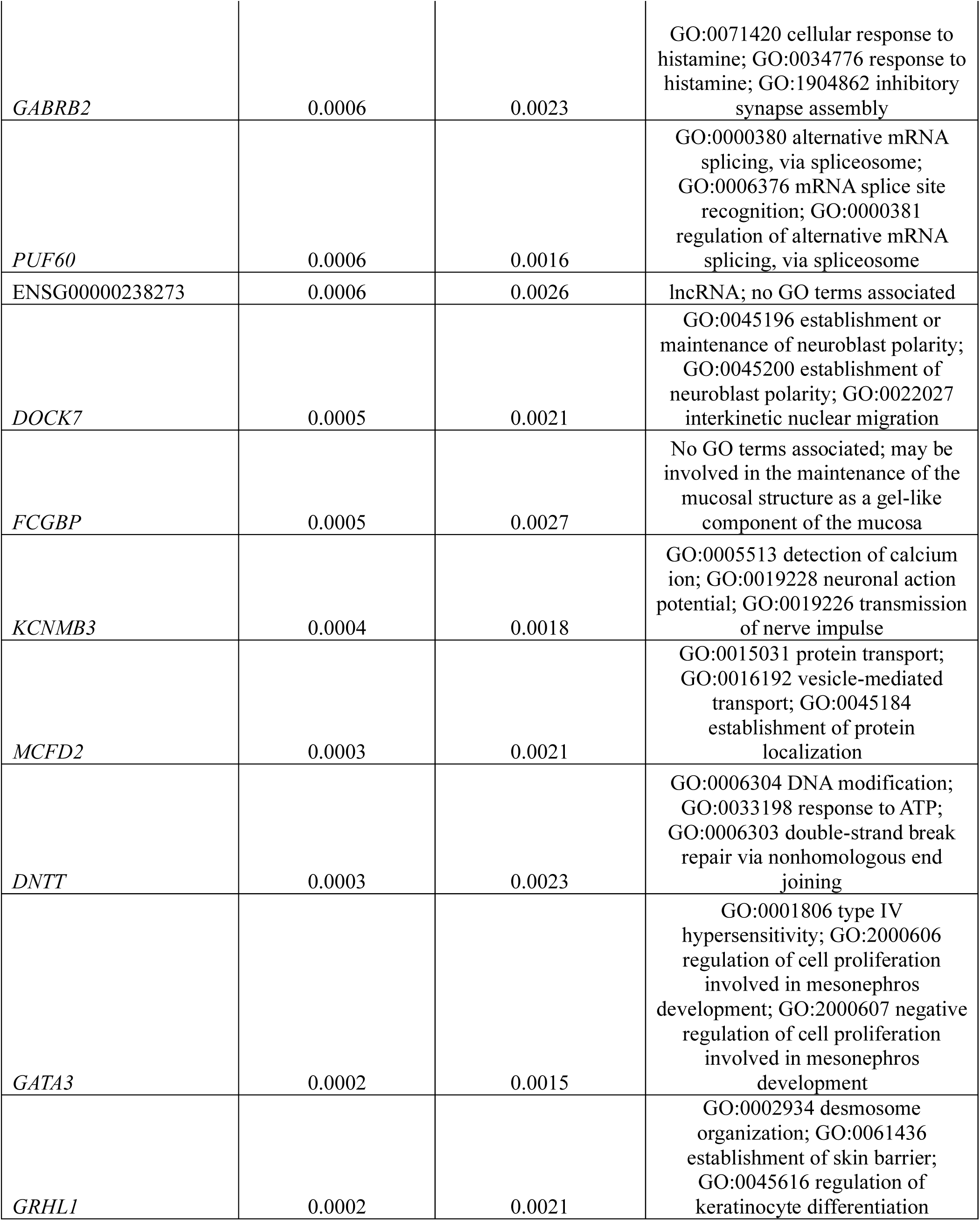
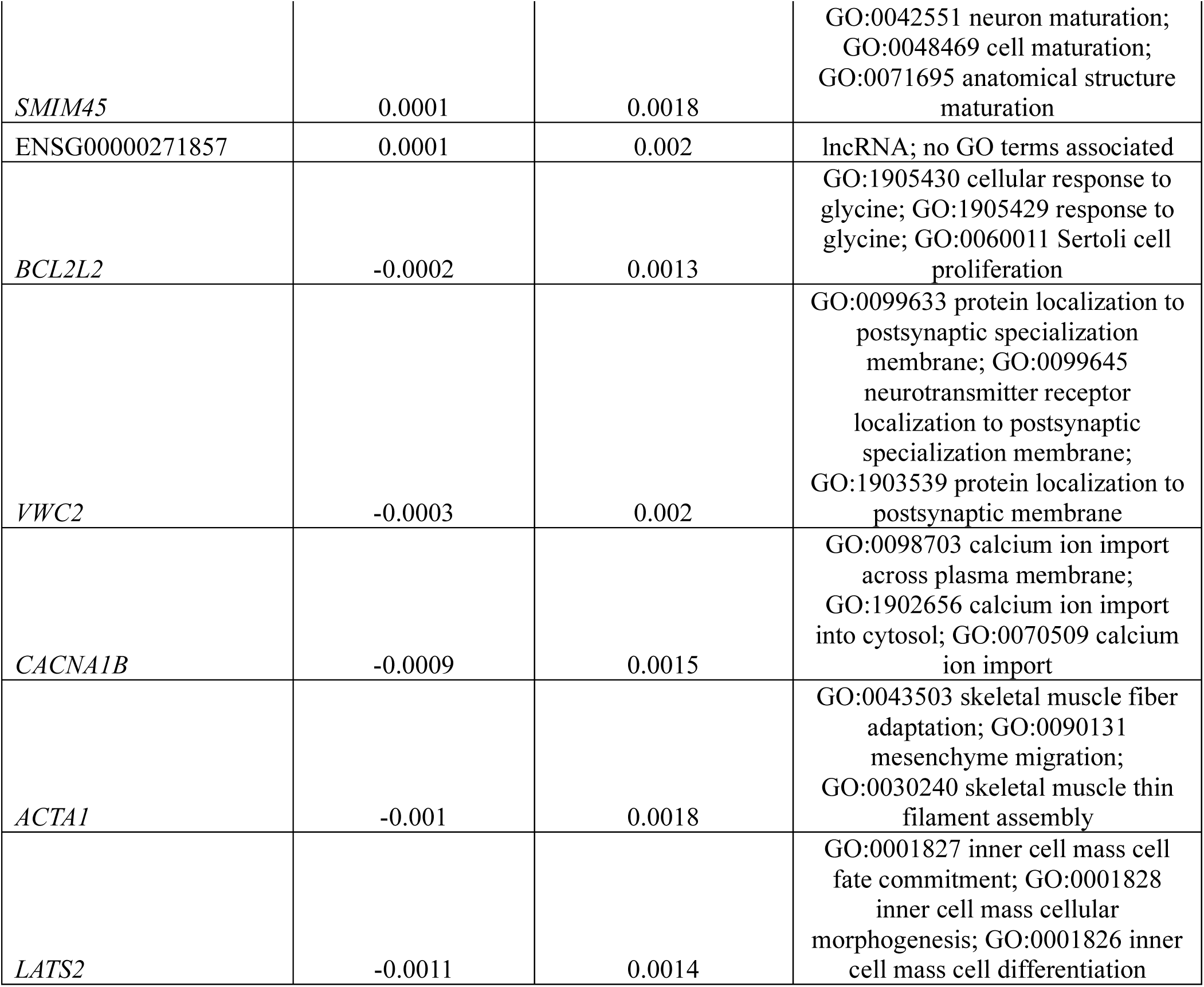
Random Survival Forest Variable Importances for OS.

**Table 7.**
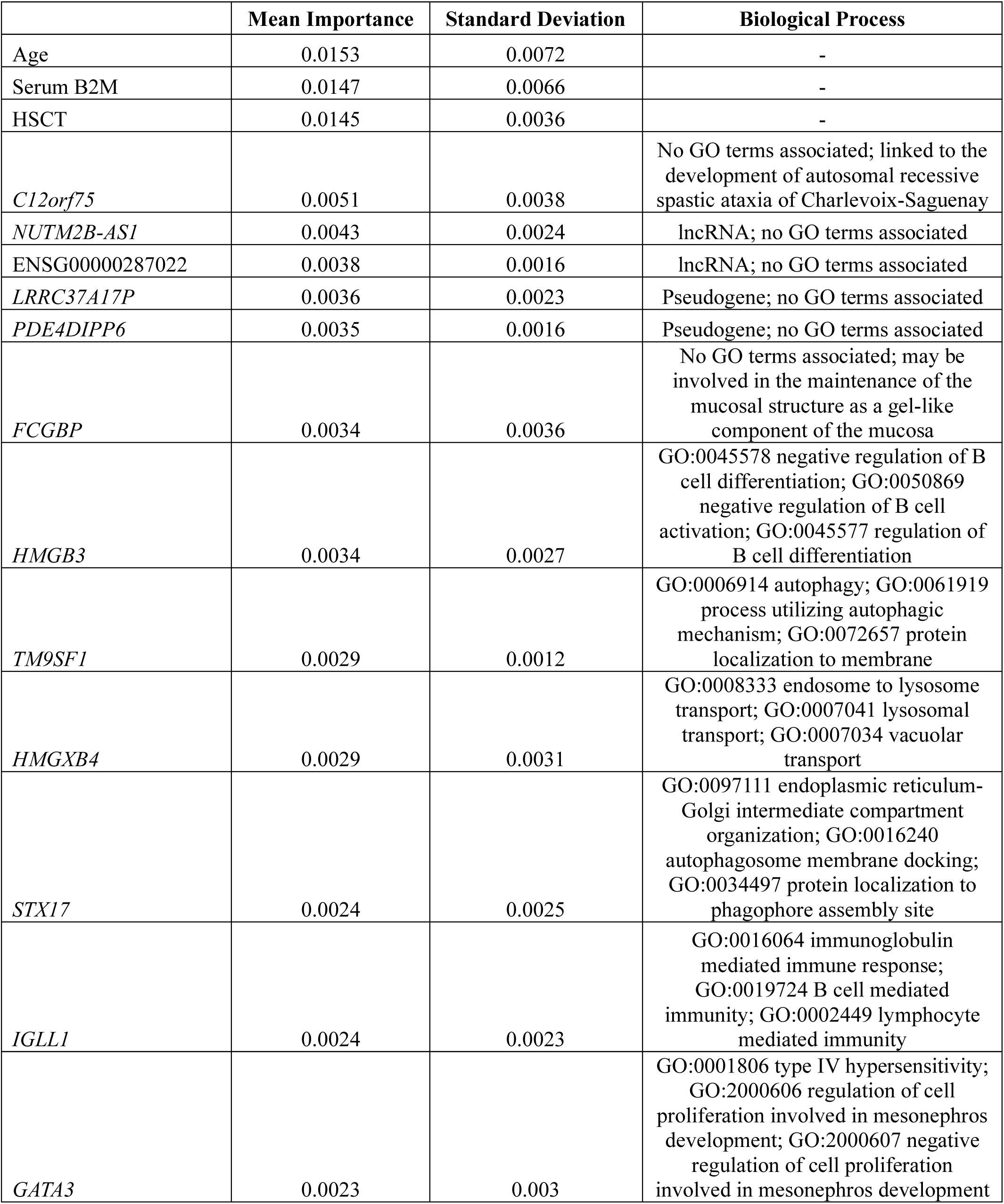
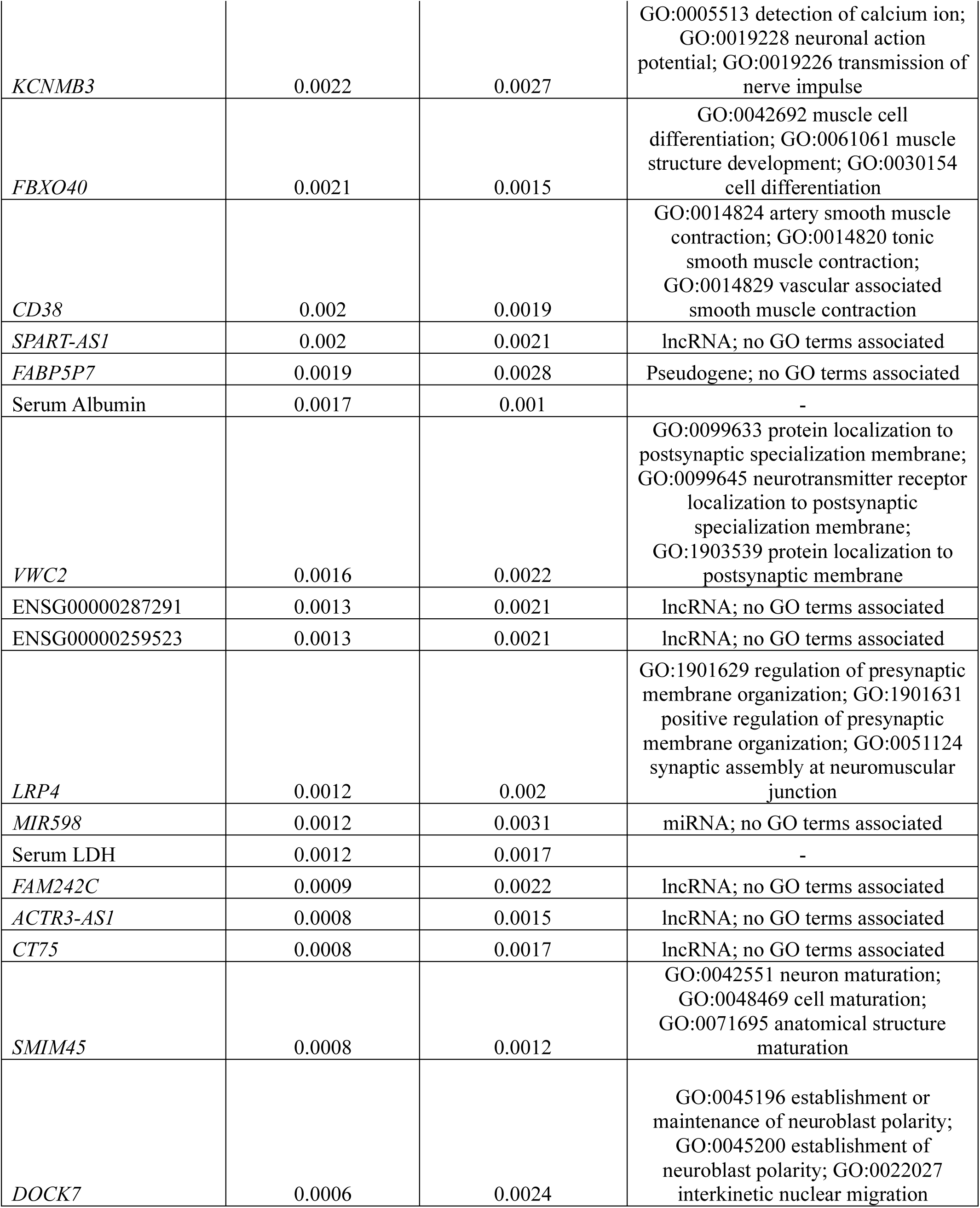
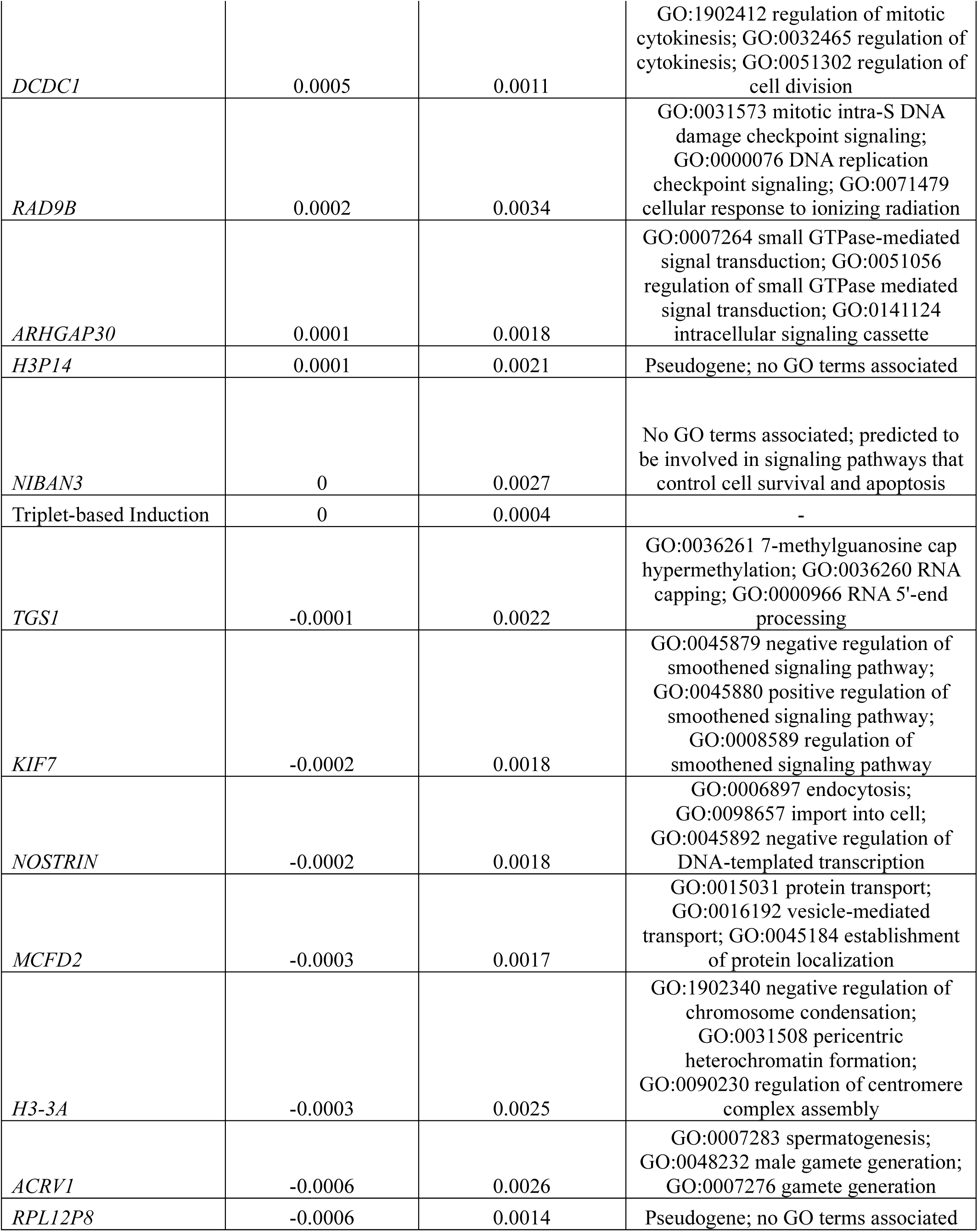
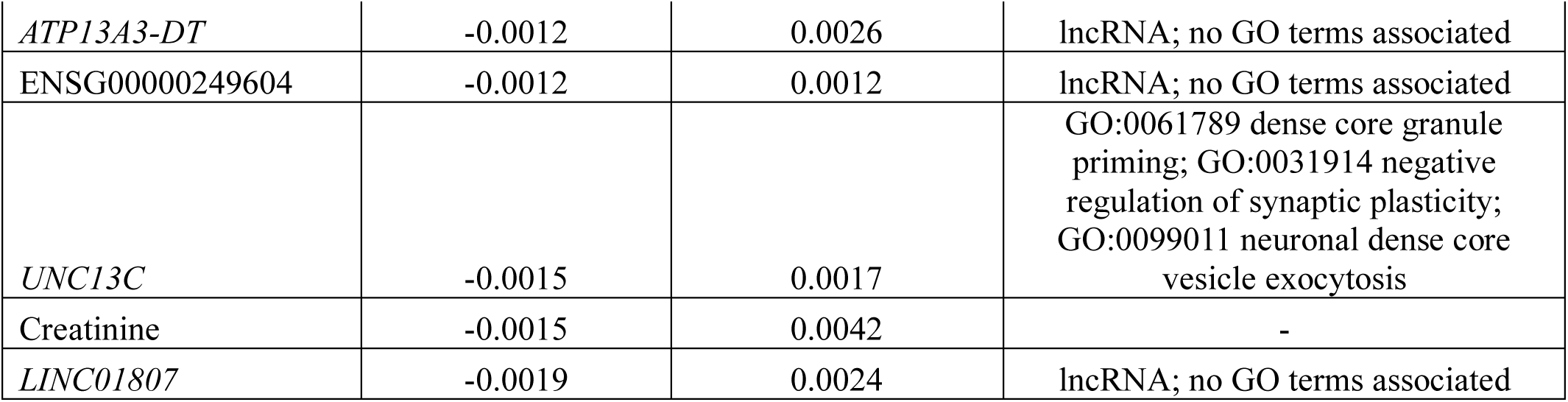
Gradient Boosted Variable Importances for OS.

|  | Mean Importance | Standard Deviation | Biological Process |
| --- | --- | --- | --- |
| Age | 0.0153 | 0.0072 | - |
| Serum B2M | 0.0147 | 0.0066 | - |
| HSCT | 0.0145 | 0.0036 | - |
| *C12orf75* | 0.0051 | 0.0038 | No GO terms associated; linked to the development of autosomal recessive spastic ataxia of Charlevoix-Saguenay |
| *NUTM2B-AS1* | 0.0043 | 0.0024 | lncRNA; no GO terms associated |
| ENSG00000287022 | 0.0038 | 0.0016 | lncRNA; no GO terms associated |
| *LRRC37A17P* | 0.0036 | 0.0023 | Pseudogene; no GO terms associated |
| *PDE4DIPP6* | 0.0035 | 0.0016 | Pseudogene; no GO terms associated |
| *FCGBP* | 0.0034 | 0.0036 | No GO terms associated; may be involved in the maintenance of the mucosal structure as a gel-like component of the mucosa |
| *HMGB3* | 0.0034 | 0.0027 | GO:0045578 negative regulation of B cell differentiation; GO:0050869 negative regulation of B cell activation; GO:0045577 regulation of B cell differentiation |
| *TM9SF1* | 0.0029 | 0.0012 | GO:0006914 autophagy; GO:0061919 process utilizing autophagic mechanism; GO:0072657 protein localization to membrane |
| *HMGXB4* | 0.0029 | 0.0031 | GO:0008333 endosome to lysosome transport; GO:0007041 lysosomal transport; GO:0007034 vacuolar transport |
| *STX17* | 0.0024 | 0.0025 | GO:0097111 endoplasmic reticulum-Golgi intermediate compartment organization; GO:0016240 autophagosome membrane docking; GO:0034497 protein localization to phagophore assembly site |
| *IGLL1* | 0.0024 | 0.0023 | GO:0016064 immunoglobulin mediated immune response; GO:0019724 B cell mediated immunity; GO:0002449 lymphocyte mediated immunity |
| *GATA3* | 0.0023 | 0.003 | GO:0001806 type IV hypersensitivity; GO:2000606 regulation of cell proliferation involved in mesonephros development; GO:2000607 negative regulation of cell proliferation involved in mesonephros development |
| *KCNMB3* | 0.0022 | 0.0027 | GO:0005513 detection of calcium ion;<br>GO:0019228 neuronal action<br>potential; GO:0019226 transmission of<br>nerve impulse |
| *FBXO40* | 0.0021 | 0.0015 | GO:0042692 muscle cell<br>differentiation; GO:0061061 muscle<br>structure development; GO:0030154<br>cell differentiation |
| *CD38* | 0.002 | 0.0019 | GO:0014824 artery smooth muscle<br>contraction; GO:0014820 tonic<br>smooth muscle contraction;<br>GO:0014829 vascular associated<br>smooth muscle contraction |
| *SPART-AS1* | 0.002 | 0.0021 | lncRNA; no GO terms associated |
| *FABP5P7* | 0.0019 | 0.0028 | Pseudogene; no GO terms associated |
| Serum Albumin | 0.0017 | 0.001 | - |
| *VWC2* | 0.0016 | 0.0022 | GO:0099633 protein localization to<br>postsynaptic specialization membrane;<br>GO:0099645 neurotransmitter receptor<br>localization to postsynaptic<br>specialization membrane;<br>GO:1903539 protein localization to<br>postsynaptic membrane |
| ENSG00000287291 | 0.0013 | 0.0021 | lncRNA; no GO terms associated |
| ENSG00000259523 | 0.0013 | 0.0021 | lncRNA; no GO terms associated |
| *LRP4* | 0.0012 | 0.002 | GO:1901629 regulation of presynaptic<br>membrane organization; GO:1901631<br>positive regulation of presynaptic<br>membrane organization; GO:0051124<br>synaptic assembly at neuromuscular<br>junction |
| *MIR598* | 0.0012 | 0.0031 | miRNA; no GO terms associated |
| Serum LDH | 0.0012 | 0.0017 | - |
| *FAM242C* | 0.0009 | 0.0022 | lncRNA; no GO terms associated |
| *ACTR3-AS1* | 0.0008 | 0.0015 | lncRNA; no GO terms associated |
| *CT75* | 0.0008 | 0.0017 | lncRNA; no GO terms associated |
| *SMIM45* | 0.0008 | 0.0012 | GO:0042551 neuron maturation;<br>GO:0048469 cell maturation;<br>GO:0071695 anatomical structure<br>maturation |
| *DOCK7* | 0.0006 | 0.0024 | GO:0045196 establishment or<br>maintenance of neuroblast polarity;<br>GO:0045200 establishment of<br>neuroblast polarity; GO:0022027<br>interkinetic nuclear migration |
| *DCDC1* | 0.0005 | 0.0011 | GO:1902412 regulation of mitotic cytokinesis; GO:0032465 regulation of cytokinesis; GO:0051302 regulation of cell division |
| *RAD9B* | 0.0002 | 0.0034 | GO:0031573 mitotic intra-S DNA damage checkpoint signaling; GO:0000076 DNA replication checkpoint signaling; GO:0071479 cellular response to ionizing radiation |
| *ARHGAP30* | 0.0001 | 0.0018 | GO:0007264 small GTPase-mediated signal transduction; GO:0051056 regulation of small GTPase mediated signal transduction; GO:0141124 intracellular signaling cassette |
| *H3P14* | 0.0001 | 0.0021 | Pseudogene; no GO terms associated |
| *NIBAN3* | 0 | 0.0027 | No GO terms associated; predicted to be involved in signaling pathways that control cell survival and apoptosis |
| Triplet-based Induction | 0 | 0.0004 | - |
| *TGS1* | -0.0001 | 0.0022 | GO:0036261 7-methylguanosine cap hypermethylation; GO:0036260 RNA capping; GO:0000966 RNA 5'-end processing |
| *KIF7* | -0.0002 | 0.0018 | GO:0045879 negative regulation of smoothened signaling pathway; GO:0045880 positive regulation of smoothened signaling pathway; GO:0008589 regulation of smoothened signaling pathway |
| *NOSTRIN* | -0.0002 | 0.0018 | GO:0006897 endocytosis; GO:0098657 import into cell; GO:0045892 negative regulation of DNA-templated transcription |
| *MCFD2* | -0.0003 | 0.0017 | GO:0015031 protein transport; GO:0016192 vesicle-mediated transport; GO:0045184 establishment of protein localization |
| *H3-3A* | -0.0003 | 0.0025 | GO:1902340 negative regulation of chromosome condensation; GO:0031508 pericentric heterochromatin formation; GO:0090230 regulation of centromere complex assembly |
| *ACRV1* | -0.0006 | 0.0026 | GO:0007283 spermatogenesis; GO:0048232 male gamete generation; GO:0007276 gamete generation |
| *RPL12P8* | -0.0006 | 0.0014 | Pseudogene; no GO terms associated |
| *ATP13A3-DT* | -0.0012 | 0.0026 | lncRNA; no GO terms associated |
| ENSG00000249604 | -0.0012 | 0.0012 | lncRNA; no GO terms associated |
| *UNC13C* | -0.0015 | 0.0017 | GO:0061789 dense core granule priming; GO:0031914 negative regulation of synaptic plasticity; GO:0099011 neuronal dense core vesicle exocytosis |
| Creatinine | -0.0015 | 0.0042 | - |
| *LINC01807* | -0.0019 | 0.0024 | lncRNA; no GO terms associated |

**Table 8.**
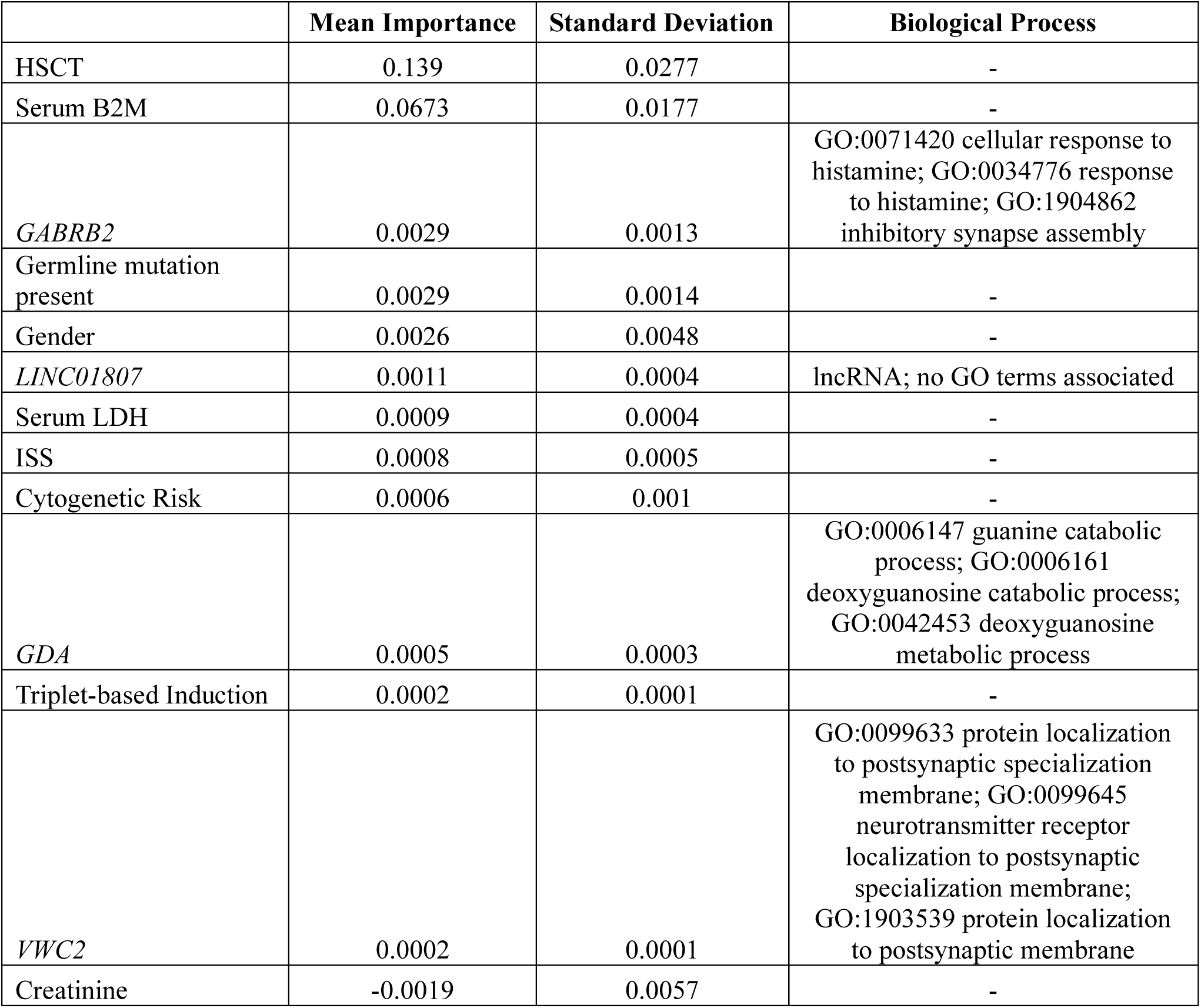
Component-wise Gradient Boosted Variable Importances for OS.

## DISCUSSION

Our study demonstrates the viability of leveraging RNA-sequencing, clinical data, and biochemical data to predict patient prognosis in multiple myeloma (MM) using various machine learning techniques. The Random Survival Forest (RSF), Gradient Boosted (GB), and Component-wise Gradient Boosted (CGB) models were developed and optimized for predicting progression-free survival (PFS) and overall survival (OS). Among these, the RSF and GB models showed the highest performance for predicting both PFS and OS. Significant predictors for PFS across models included clinical features like HSCT status, serum β2-microglobulin levels, and gene expression levels of *C12orf75* and ENSG00000256006, particularly for the RSF and GB models **(Figure 3)**. The most significant gene expression RSF and GB features for predicting PFS included long non-coding RNA (lncRNA) transcripts and pseudogenes. Both have been shown to regulate gene expression networks via competing endogenous RNA (ceRNA) networks and potentially be a prognostic biomarker for patients with colorectal cancer, renal cell carcinoma, and lung adenocarcinoma (Li et al., 2022; Stewart et al., 2019; Chen et al., 2018). Pan-cancer analysis of pseudogene expression, when combined with clinical variables, suggested extensive and robust concordance with other molecular subtyping methods and risk stratification techniques for patients with kidney cancer (Han et al., 2014).

Significant predictors for OS included HSCT status, serum β2-microglobulin levels, age, and, to a lesser extent, ISS. Gene expression levels of *NUTM2B-AS1*, ENSG00000287022, *LRRC37A17P*, and *PDE4DIPP6* were significant features for the RSF and GB models (**Figure 3)**. These are similarly noteworthy as they fall within the broad classes of lncRNAs and pseudogenes, which were also seen as significantly predictive of PFS. However, despite these similarities, heterogeneity remains in the gene expression features most relevant for predicting patient prognosis, particularly when considering the biological processes of each outcome’s coding transcripts. The gene ontology and pathway enrichment analyses implicated pathways related to cell division, protein localization, and cancer, among others, for PFS. Pathways related to *Wnt* signaling, RNA splicing, and PPARA activation were predictive for OS.

Clinical features, however, were shared and generally similar in importance between the RSF and GB models **(Figure 5)**. The CGB model was an outlier, most likely because it primarily bases predictions on clinical features and gene expression profiles not found in the other models **(Figure 4)**. This may account for the decreased performance of the CGB model compared to the RSF and GB models and demonstrates the value of gene expression features as prognostic biomarkers for patients with MM.

**Figure 4.**
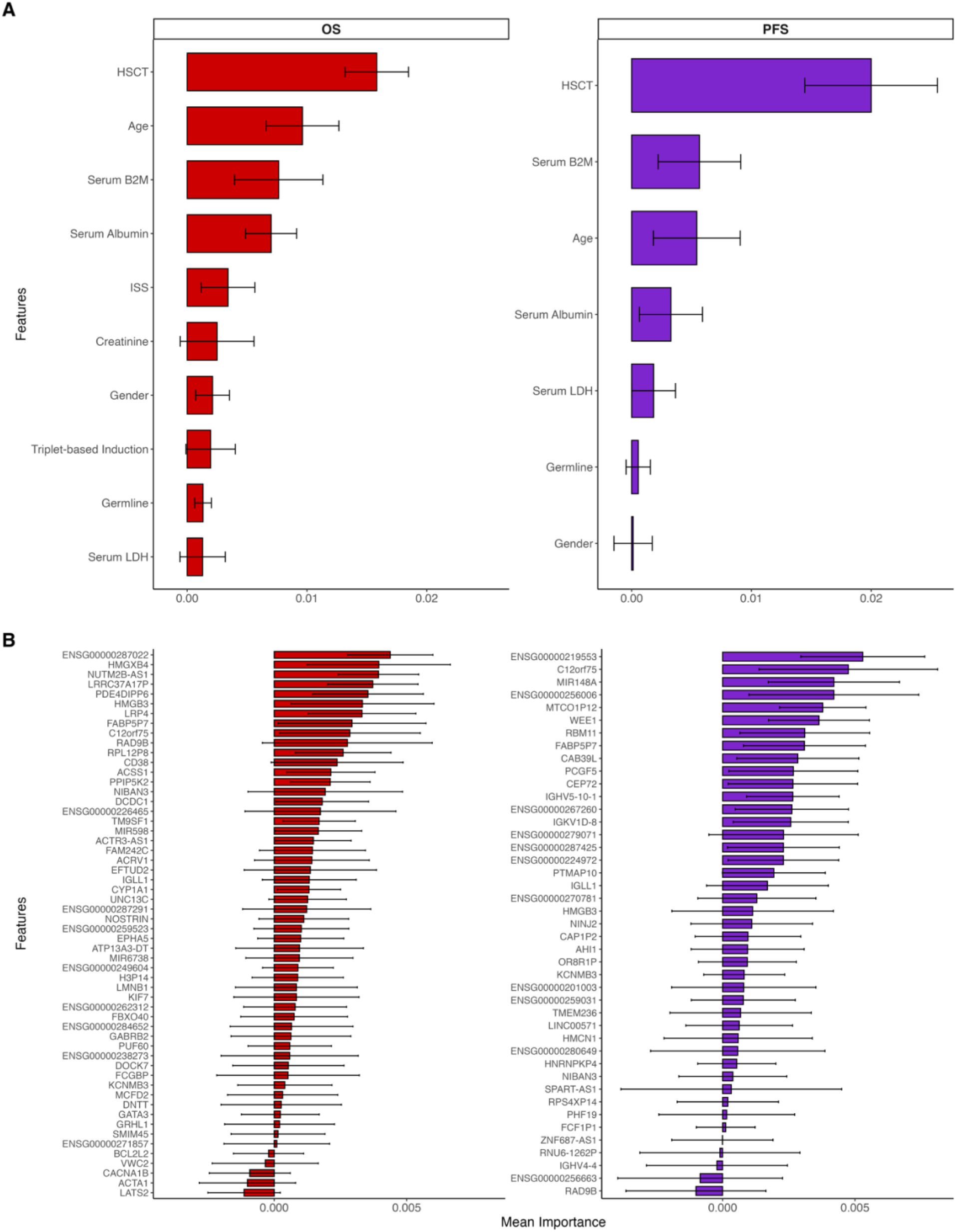
Clinical and Gene Expression Feature Importances for the Random Survival Forest Model **A** Clinical feature importances faceted by outcome. The error bars represent mean importance plus or minus the associated standard deviation. **B** Gene expression feature importances faceted by outcome.

**Figure 5.**
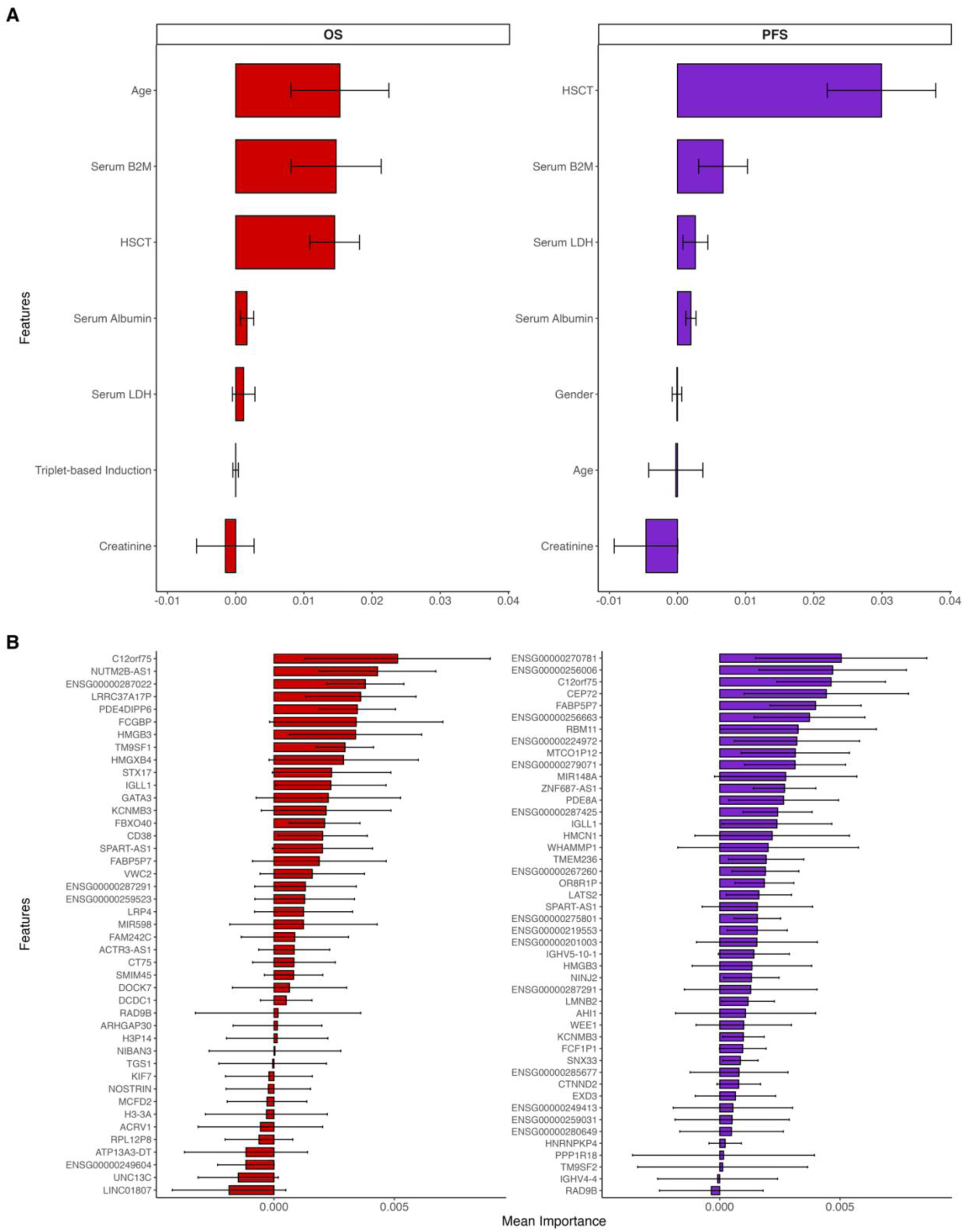
Clinical and Gene Expression Feature Importances for the Gradient Boosted Model **A** Clinical feature importances faceted by outcome. The error bars represent mean importance plus or minus the associated standard deviation. **B** Gene expression feature importances faceted by outcome.

In terms of overall performance for predicting OS, our RSF and GB models compare favorably to examples published in the literature, all of which achieved concordance indices ranging from 0.64 to 0.78 on internally and externally validated testing datasets (Hummel et al., 2024, Orguiera et al., 2021; Terebelo et al., 2019; Perrot et al., 2019). In comparison, our study expands upon prior MM machine learning models by including PFS as an outcome and by adding additional clinical and biochemical features like germline mutational status (Orgueira et al., 2021). Germline mutational status, while previously known to be associated with improved PFS, can also predict OS in our machine learning model (Thibaud et al., 2024).

While some features were shared from prior examinations of the CoMMpass dataset (serum β2-microglobulin levels, age, ISS, *WEE1* gene expression), many were unique to our models, which demonstrates the value of different machine learning models in uncovering novel genetic and clinical biomarkers that may provide additional insights into the understanding of multiple myeloma biology and clinical prognosis (Orgueira et al., 2021). Intriguingly, cytogenetic risk failed to predict for PFS or OS in our highest-performing models and the IAC- 50 model developed by Orgueira and colleagues (Orgueira et al., 2021). This likely stems from redundant and noisy relationships with other features in the data. This underscores the value of advanced feature selection techniques, like mRMR, for enhancing model performance and allowing for more downstream iteration and fine-tuning.

There are several limitations to our study. Although relatively large, the sample size remains relatively small for generating a machine learning model. Our findings will require external validation across multiple cohorts to confirm the findings’ generalizability. This limitation will likely require multi-center collaboration and data harmonization—a true testament to the work of the MMRF staff and research team in generating the CoMMpass dataset. While imputation was used to handle missing data, it might introduce biases that could affect the results (Woods et al., 2023). Additionally, our models may suffer from overfitting, despite efforts to mitigate this through Monte-Carlo cross-validation and pruning in the optimization process.

Future research should focus on independent validation using external cohorts to ensure the robustness and applicability of our models. Exploring other advanced machine learning or deep learning techniques using multi-modal data could also enhance prediction accuracy. Expanding the analysis to include data related to structural variants may also provide more comprehensive insights into the genetic underpinnings of MM prognosis and enable more accurate and precise prognostication.

## Data Availability

All data produced in the present study are available upon reasonable request to the authors.

https://research.themmrf.org/

https://www.ncbi.nlm.nih.gov/projects/gap/cgi-bin/study.cgi?study_id=phs000748.v7.p4).

## Notes

### Author Declarations

Clinical, biochemical, survival, and RNAseq data was acquired from the semi-public Multiple Myeloma Research Foundation (MMRF) Research Gateway (https://research.themmrf.org/). Raw whole exome sequencing data was downloaded from the Database of Genotype and Phenotype (#phs000748.v7.p4, project #34982, https://www.ncbi.nlm.nih.gov/projects/gap/cgi-bin/study.cgi?study_id=phs000748.v7.p4).

